# Research themes of family and community physicians in Brazil

**DOI:** 10.1101/2021.12.22.21268269

**Authors:** Leonardo Ferreira Fontenelle, Stephani Vogt Rossi, Miguel Henrique Moraes de Oliveira, Diego José Brandão, Thiago Dias Sarti

**Affiliations:** Universidade Vila Velha (UVV), Medical School. Vila Velha (ES), Brazil; Universidade Vila Velha (UVV), Medical School. Vila Velha, ES, Brazil; Universidade Federal do Espírito Santo (UFES), Center of Health Sciences, Department of Social Medicine. Vitória (ES), Brazil

**Keywords:** Physicians, Family, Brazil, Bibliometrics, Biomedical Research, Cluster Analysis, Education, Graduate

## Abstract

Family and community medicine is a specialty dedicated to primary care, the cornerstone of effective health systems. Research capacity in primary care varies worldwide, and bibliographic databases such as MEDLINE, Scopus and Web of Science do not index most primary care research coming from Latin America. Our objective was to investigate the research themes of family and community physicians in Brazil, and to correlate the articles’ research themes with their authors’ trajectories in postgraduate education. For that, we compiled a national list of family and community physicians, retrieved their curricula from the Lattes Platform, compiled a list of journal articles, and obtained their keywords from LILACS and MEDLINE. Treating journal articles and their keywords as the two node types in a bipartite network, we derived research themes using the dual-projection algorithm, combining the Leiden algorithm with hierarchical clustering. We found two research themes to be the largest, most developed, and most central ones: human health and primary care. Authors with a master’s or PhD in collective health (public health, epidemiology, and social sciences and humanities in health) were as likely as those with no postgraduate degree to publish articles on primary care. On the other hand, authors with a postgraduate degree in medicine were more likely to publish articles on human health. After discussing the findings in light of previous research and methodological aspects, we conclude there’s a relative divide between primary care and clinical research, and the highlight policy implications.

## Introduction

Family and community physicians, also called family doctors or general practitioners, provide comprehensive, patient-centered healthcare to individuals of all age groups, regardless of the affected organs (including mental health) (Arya et al. 2017; Freeman and McWhinney 2016; Gupta et al. 2021; Romano 2008). They became a separate specialty during the second half of the 20^th^ century, with the increased recognition of primary health care as a cornerstone of effective health systems (Bonet 2014; Ceitlin 2006; Falk 2004; Freeman and McWhinney 2016; Gupta et al. 2021; Pires-Alves and Paiva 2021; Ponka et al. 2015; Rodrigues 2007). In primary health care, patients are enrolled in healthcare services where they have timely access to comprehensive healthcare (Starfield et al. 2005). Because different stakeholders emphasize different aspects of primary health care, the expression “primary care” is often preferred for the more clinical aspects (Gupta et al. 2021; Muldoon et al. 2006; Ponka et al. 2015). Here, though, we use both terms interchangeably.

Primary care research embraces diverse research methods and paradigms, and encourages collaboration with disciplines such as public health and behavioral science (Freeman 2012; Goodyear-Smith et al. 2016; McWhinney 1996). Besides clinical practice, primary care research comprises research in health services, health systems, medical education, and research methodology (Beasley et al. 2004, 2007). Four decades ago, English-language journals of family and community medicine published more research *about* primary care than *in* primary care (Freeman 2012), and two decades ago McWhinney (2001a, 2001b) lamented family and community physicians did little clinical research. Since then, primary care research acquired a clinical focus – at least, in Western countries. Looking on the most-often cited papers in English-language journals, Freeman (2012) derived six research themes: clinical issues, epidemiology in family practice, patient experiences, research methods, physician issues, and conceptual. More recently, López-Torres Hidalgo (2014, 2020) surveyed primary care research in Spain, and found that two-thirds of it focused on clinical aspects.

Historically, research capacity in primary care follows the academic presence of family and community medicine, which in turn follows its recognition as a medical specialty (Beasley et al. 2004; Goodyear-Smith and Mash 2016). To this day, there is wide international variation in the research capacity of family and community medicine. In the UK and the Netherlands, for example, most primary care is provided by family and community physicians, and most or all universities have departments dedicated to primary care (van der Zee et al. 2003). These countries happen to be some of the top performers in primary care research (Glanville et al. 2011). In many countries, though, family and community medicine is still struggling for recognition (Arya et al. 2017), and primary care research is inexpressive.

Brazil is an intermediate case. Together with China, India and South Africa, the country is home to a large share of the world’s population, and is committed to strengthening primary health care, even though it is not quite there yet (Mash et al. 2015). In Brazil, family and community medicine is formally recognized as a medical specialty, and there is an increasing number of medical residency programs, but most medical providers in primary care are not family and community physicians (Mash et al. 2015). Furthermore, neither family and community medicine, nor primary health care is officially recognized as a distinct research area for the purposes of funding research or assessing postgraduate programs (masters’ and doctorates) (Wenceslau et al. 2020). As a consequence, most family and community physicians in Brazil earn their postgraduate degrees in collective health (Fontenelle et al. 2020), a discipline in Latin America comprising public health, epidemiology, and humanities and social sciences in health (Osmo and Schraiber 2015; Vieira-da-Silva and Pinell 2014). Family and community physicians in Brazil also publish predominantly in collective health journals (Fontenelle, Oliveira, et al. 2021), or “public health” journals, as the US National Library of Medicine categorizes them.

As in other developing countries (Domínguez et al. 2016; Escobar et al. 2018; Ortega et al. 2016; Sparks and Gupta 2004), primary care research in Brazil has been hindered by low access to funding and postgraduate education, and obstacles to practice-based research. As a consequence, primary care research in Brazil has been described as low-volume, low quality, of little consequence, relying too much on descriptive studies, and being published in Portuguese in journals not even indexed in LILACS, the Latin American and Caribbean Health Sciences Literature database (Almeida et al. 2016; Harzheim et al. 2005).

This dire situation might be changing, though. MacAulley (2011) argues “Brazilians have the potential to be future leaders in primary care”, based on the scale and youth of the Brazilian Conference on Family and Community Medicine. Scale and youth indeed: in late 2018, half the family and community physicians in Brazil had entered the specialty in the preceding 10 years (Fontenelle et al. 2020). Furthermore, Brazilian family and community physicians increasingly publish in journals from other countries, suggesting their research is increasingly of international relevance (Fontenelle, Oliveira, et al. 2021).

Studying the making of science in developing countries such as Brazil might complement previous studies focusing on developed countries. Indeed, Brazil has been described as representative of, and a leader in, Latin America (Glänzel et al. 2006). Furthermore, the health sciences have been found to be the one of the largest research areas in Brazil, and one of the most internationalized ones (Barata et al. 2014), while having bibliometric indicators similar to the national average of all research areas (Grácio and Oliveira 2014).

Such large-scale studies necessarily rely on international databases such as Scopus and the Web of Science. However, even though Brazilian journals are increasingly indexed in such databases (Leta et al. 2013), progress is heterogeneous. As we write this paper, there are no journals from Latin America in the “primary health care” category of Web of Science, the “family practice” subject area of Scopus, or the “primary health care” broad subject term of MEDLINE (except for one internal medicine journal from Colombia).

In other words, family and community medicine in Latin America is poorly represented in large-scale studies of scientific publication. Fortunately, Brazil’s Lattes Platform provides an excellent data source (Lane 2010). Created in 1999, it is the Brazilian information system on science, technology and innovation. Curricula in the Lattes Platform must be kept reasonably complete and up to date, because they are used for decisions on research funding and on recruitment, promotion and tenure. Furthermore, these curricula must be reasonably honest because they are publicly available, and researchers are accountable for the information they provide. Indeed, the Lattes Platform has been used in previous scientometric research (Leite et al. 2011; Mena-Chalco et al. 2014; Sidone et al. 2016).

In this research paper, we make use of the Lattes Platform to investigate the research themes of family and community physicians in Brazil, and correlate these themes with their authors’ trajectories in postgraduate education.

## Methods

This was an exploratory study, using administrative data from multiple sources, as part of the *Trajetórias MFC* project (Fontenelle et al. 2020; Fontenelle, Oliveira, et al. 2021). The project was approved by the Research Ethics Committee of Universidade Vila Velha (report n. 3.033.911). The anonymized dataset is open (Fontenelle, Rossi, et al. 2021b), and the underlying datasets are available upon request (Fontenelle et al. 2019; Fontenelle, Rossi, et al. 2021a).

### Data sources

In Brazil, there are two ways for a physician to be recognized as a specialist: to conclude postgraduate training in the specialty (medical residency) or to pass a certification exam. Accordingly, a nationwide list of family and community physicians as of late 2018 was compiled from two data sources, as described in Fontenelle et al. (2020): the information system for medical residency (SisCNRM) and a list of certified physicians, provided by the Brazilian Society of Family and Community Medicine (*Sociedade Brasileira de Medicina de Família e Comunidade*, SBMFC). The physicians’ genders were inferred from their first name, using data from the 2010 Brazilian Census. Data on their *stricto sensu* postgraduate education (masters’ and doctorates) were obtained from their curricula in the Lattes Platform and the Sucupira Platform, which is the Brazilian information system on postgraduate programs (Siqueira 2019).

The list of journal articles published by the family and community physicians was obtained in August 2019 from their curricula in the Lattes Platform, as described in Fontenelle et al. (2021). After identifying them in the curricula, we looked the journal articles up in Cross-Ref, MEDLINE (through PubMed) and LILACS (through Pan-American Health Organization’s Virtual Health Library [VHL]), during late 2019 through early 2020. Furthermore, we looked journals up in the catalog of the US National Library of Medicine (NLM), and settled any doubts by looking up in the ISSN International Center ROAD (Directory of Open Access scholarly Resources) and the VHL Serials in Health Sciences (SeCS). The data were verified for internal inconsistencies throughout the entire process, often resulting in the deletion of a duplicated entry or a minor adjustment of citation data, but we defaulted to trusting our data sources.

### Measures

Unless otherwise specified, all results refer to articles published during or after the year of specialization in family and community medicine. For physicians with both a medical residency and certification, we considered the earliest of the two years. For descriptive purposes, publication dates were grouped in five-year periods, with the first period having an unbounded beginning (any year, up to 1998). Physicians were considered female when their first name had ≥ 50% probability of belonging to a woman, and male otherwise. We categorized the knowledge area of the postgraduate degrees (masters’ and PhDs) as “none” (no postgraduate degree, or no degree yet), “medicine”, “collective health” and “other”.

Articles were weighted by harmonic counting of authorship, with the last author receiving equal weight as the first one (Hagen 2008). Harmonic counting weights authorship according to the author’s position in the byline, and closely fits authorship credit perceived by peers (Hagen 2010, 2013). In our study, articles weighted less than 1 if not all authors were family and community physicians. If an author specialized only after publishing an article, we did not count this as an authorship by a family and community physician.

The articles’ subject was described using the US NLM’s Medical Subject Headings (MeSH) (Rogers 1963), for articles found in MEDLINE, or Health Sciences Descriptors (DeCS), for articles found in LILACS. DeCS is a superset of MeSH, with the addition of contextually relevant subject headings. We opted for DeCS when an article was indexed in both MEDLINE and LILACS, and excluded articles not indexed in either. In this paper, we use “headings”, “descriptors” and “keywords” interchangeably.

### Data analysis

We analyzed our data as an undirected bipartite network, with articles and keywords being the two node types, and edges being the occurrence of keywords in articles. For each article, the weight of the edges to the corresponding keywords was the inverse of the number of keywords. As a consequence, each article had a strength centrality (Barrat et al. 2004) of 1. The weight of the keywords was the weight of their edges, multiplied by the weight of the corresponding articles (as in Measures), then summed. As a consequence, the sum of the keyword weights was the same as the sum of the article weights.

The articles and keywords were clustered into research themes using the dual-projection algorithm (Arthur 2020; Everett and Borgatti 2013; Melamed 2014). The first step of the algorithm consists in obtaining a “top” (keyword co-occurrence) and a “bottom” (articles as nodes, shared keywords as edges) projection of the bipartite network. As in Cobo (2011), we normalized these projections so that the resulting edge weights did not depend anymore on the frequency of the corresponding keywords or articles. In our case, the edges were normalized with an improved version of the association strength (Steijn 2021). The association strength has an expected value of 1 (Eck and Waltman 2009; Steijn 2021), while the equivalence index used in Cobo (2011) has a maximum value of 1.

The second step consists in independently clustering the top and the bottom projections. Here we opted for the Leiden algorithm (Traag et al. 2019), while Arthur (2020) used the Louvain algorithm (Blondel et al. 2008), Melamed (2014) used the walktrap algorithm (Pons and Latapy 2006) and Everett and Borgatti (2013) used the singular value decomposition (Trefethen and Bau III 1997). The resolution parameter of the Leiden algorithm was tuned to provide a reasonable number of reasonably-sizes clusters.

The third and final step of the dual-projection algorithm consists of creating an aggregate bipartite network from the two sets of clusters (the top and bottom ones) and then running this aggregate network through hierarchical clustering. Following Newman (2004), the hierarchical clustering algorithm chose edges for removal by dividing the unweighted edge betweenness by the edge weights. For consistency with the choice of the Leiden algorithm over the Louvain one, the optimal cut of the hierarchical clustering was decided by optimizing the constant Potts model (Traag et al. 2011) of the bipartite network, instead of optimizing the modularity (Newman and Girvan 2004). The resolution parameter was tuned by inspecting the dendrogram of the hierarchical clustering, not the contents of the clusters.

The resulting research themes were named after the most frequent keyword (that is, that with the largest weight). Had we named our themes after the keywords with higher strength centrality as Cobo et al. (2011), the resulting names would clash with subject-matter expertise because our algorithm, contrary to theirs, preserves edges with large weights between rare keywords. We also used weighted frequency to select representative keywords for inclusion in co-occurrence diagrams representing the scholarly output as a whole as well as for each of the three main themes. The number of keywords in such diagrams was based on clarity of visualization.

To describe the research themes, we used the centrality and density of their keyword co-occurrence representations (Callon et al. 1991; Cobo et al. 2011), as well as the weighted number of articles. The themes’ centrality was calculated as the sum of the edges between them and other themes, and indicates their relevance to the overall scholarly output. The themes’ density was calculated as the sum of its internal edges divided by the number of keywords, and indicates their internal cohesion or development ^1^. The centrality index was divided by 100, and the density index was multiplied by 100. The research themes were also graphically described using a strategic map (Callon et al. 1991; Cobo et al. 2011), which plots the themes as bubbles with the scaled centrality and density indexes as the axes and the weighted number of articles as the bubble size.

We also described the physicians’ trajectories in postgraduate education leading to each of the largest research themes, as well as to the total scholarly output. For that purpose, we tabulated the weighted frequency of articles by knowledge area (none, medicine, collective health, other) and level (none, master’s, PhD) of the latest postgraduate degree, as well as by the physicians’ gender and mode of specialization (medical residency or certification).

We also tabulated the five-year periods of publication, to describe the time trends in publication volume.

Network analyses were conducted with packages igraph (Csardi and Nepusz 2006), development version 1.2.8, and Matrix, version 1.3-4, for the R language and environment for statistical computing, version 4.1.2, with OpenBLAS, verson 0.3.15. The network diagrams were laid out with the Fruchterman and Reingold (1991) algorithm (as implemented by the igraph package), and plotted with the ggplot2 3.3.5 (Wickham 2016) and ggrepel 0.9.1 packages for R. The color of the research themes was chosen from the colorblind-friendly palette suggested by Okabe and Ito (2002). We wrote our own implementation of the hierarchical clustering algorithm and constant Potts model to fit the bipartite nature of our data. Our code is openly available (Fontenelle 2021).

## Results

As of late 2018, Brazil had 6238 family and community physicians, of which 4065 (65.2%) had a curriculum in the Lattes Platform and 1111 (17.8%) had 4918 unique articles (1635.1 articles, weighting by authorship). Disregarding articles published before specialization, 636 (10.2%) family and community physicians had 3154 unique articles (1100.0 articles, with weighting). Further disregarding articles without keywords in MEDLINE (MeSH) or LILACS (DeCS), 432 physicians had 1904 unique articles (577.5 articles, with weighting). We further excluded one article which did not share its keywords with any other article, resulting in 1903 unique articles and 3222 unique keywords, both amounting to 576.5 with authorship weighting.

Family and community physicians accounted for 30.3% of the articles’ authorship, with coauthors presumably being colleagues in training or from other countries, specialties or professions. There were 8.8 keywords per article, on average. Figure 1 displays the co-occurrence between the most common keywords, amounting to half the weighted frequency.

**Figure 1.**
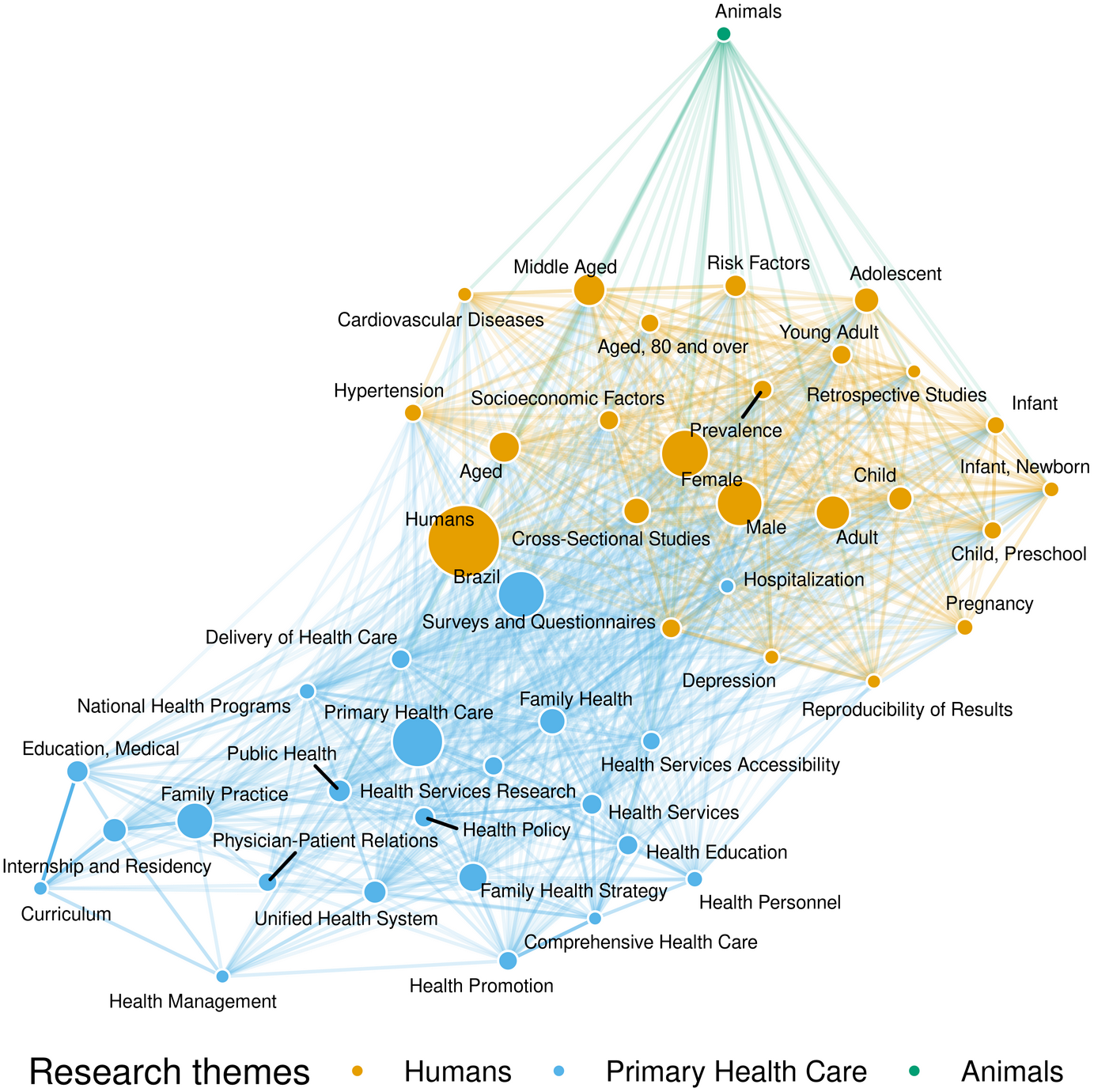
Diagram of keyword co-occurrence for the most common keywords from articles authored by family and community physicians in Brazil. Bubble areas are proportional to weighted frequency and edges’ opacity is proportional to their normalized weight.

The clustering algorithm detected 268 research themes, of which only 14 included at least two unique articles and two unique keywords (Suppl. Table 1). Three research themes included 85.5% of the articles:

1. *Humans* was the largest theme, with 271.1 (47.0%) articles and 242.7 (41.1%) keywords. The most common keywords (Suppl. Figure 1) described characteristics of human populations (eg humans, female, aged), study design (eg prevalence, surveys and questionnaires, retrospective studies) and clinical conditions (eg cardiovascular diseases, chronic diseases, HIV infections), among others (eg socioeconomic factors, psychotherapy, reproducibility of results).
2. *Primary health care* was the second largest theme, with 194.5 (33.7%) articles and 226.0 (39.2%) keywords. The most common keywords (Suppl. Figure 2) described primary care (eg primary health care, family practice, Family Health Strategy), public health (eg Unified Health System, public health, health promotion), medical education (eg medical education, internship and residency, curriculum), study methods (qualitative research) and geography (Brazil).
3. *Animals* was the third largest theme, with 27.2 (4.7%) articles and 31.5 (5.5%) keywords. The most common keywords (Suppl. Figure 3) described animals used in preclinical research (eg animals, rats, Winstar rats), clinical conditions (eg mental health, Chagas disease, complex regional pain syndrome), morphology, physiology or pathology (eg blood glucose, oxidative stress, *Trypanosoma cruzi*) or therapy (eg psychoanalysis, homeopathy, physical education).

The average article in the *humans* research theme was 27.3% authored by family and community physicians (Suppl. Table 1). This is about the overall average (30.3%), while lower than in the *primary health care* theme (45.2%) and higher than in the a*nimals* theme (17.4%).

Articles in both the *humans* (10.4) and the *animals* (12.4) research themes had more keywords than the overall average (Suppl. Table 1). On the other hand, articles in the *primary health care* theme had only 5.5 keywords on average.

The three largest research themes were also the most central ones, with *primary health care* and *animals* being more strongly connected to *humans* than among themselves (Figure 2). *Humans* and *primary health care* were the most developed (dense) themes, while *animals* was less developed than some of the smaller themes.

**Figure 2.**
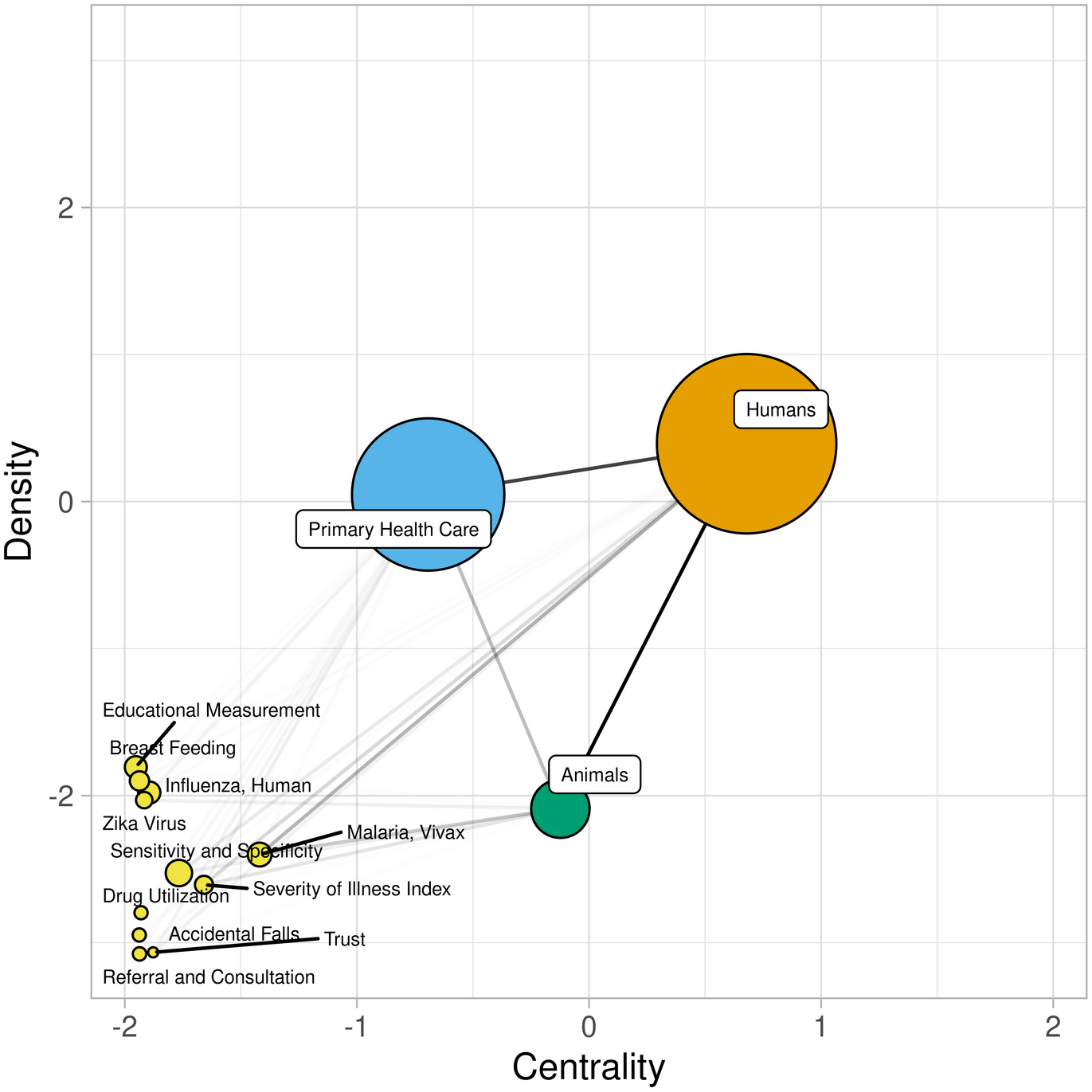
Strategic map of the research themes of family and community physicians in Brazil. Bubble areas are proportional to the weighted number of articles, and edges’ opacity is proportional to their weight.

The number of articles increased over the time periods (Figure 3, Suppl. Table 2). Before 2004, research themes *primary health care* and *animals* were even smaller in comparison to *humans*, and the smaller themes didn’t even show up. Throughout the next time periods, smaller themes started being published, and the three largest themes converged to centrality and density indexes (Figure 3) similar to those of the total time frame (Figure 2). *Primary health care* saw the largest increase from years 1999–2003 to years 2009–2013, both in relative and absolute numbers (Suppl. Table 2).

**Figure 3.**
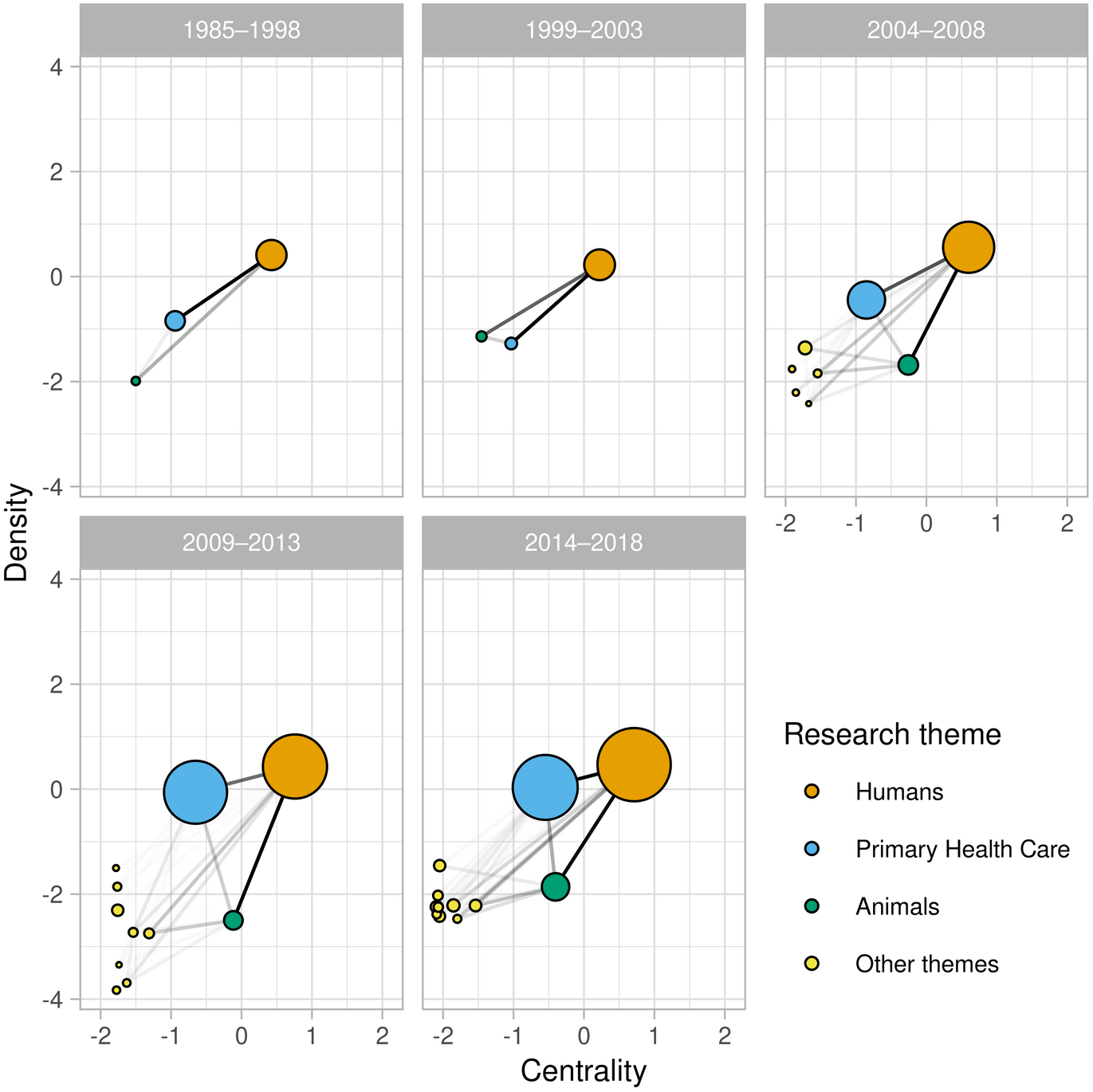
Evolution of the research themes’ strategic map over time. Bubble areas are proportional to the weighted number of articles, edges’ opacity is proportional to their weight, five-year periods refer to publication date.

Male family and community physicians accounted for most of the authorship overall (64.2%) and in research themes *humans* (64.7%) and *primary health care* (68.9%), but not in *animals* (39.3%) (Suppl. Table 2). Likewise, physicians with a medical residency accounted for most of authorship overall (70.6%), with the proportion raging from 74.0% in *humans* to 57.0% in *animals*.

Family and community physicians with a *stricto sensu* (master’s or PhD) degree in medicine or collective health accounted for two thirds of the authorship (Figure 4; Suppl. Table 2). Most authors in the *humans* research theme had a master’s or PhD in medicine (40.7%) but for the *primary health care* theme it was usually collective health (41.2%) and for *animals* it was usually other knowledge area (45.0%).

**Figure 4.**
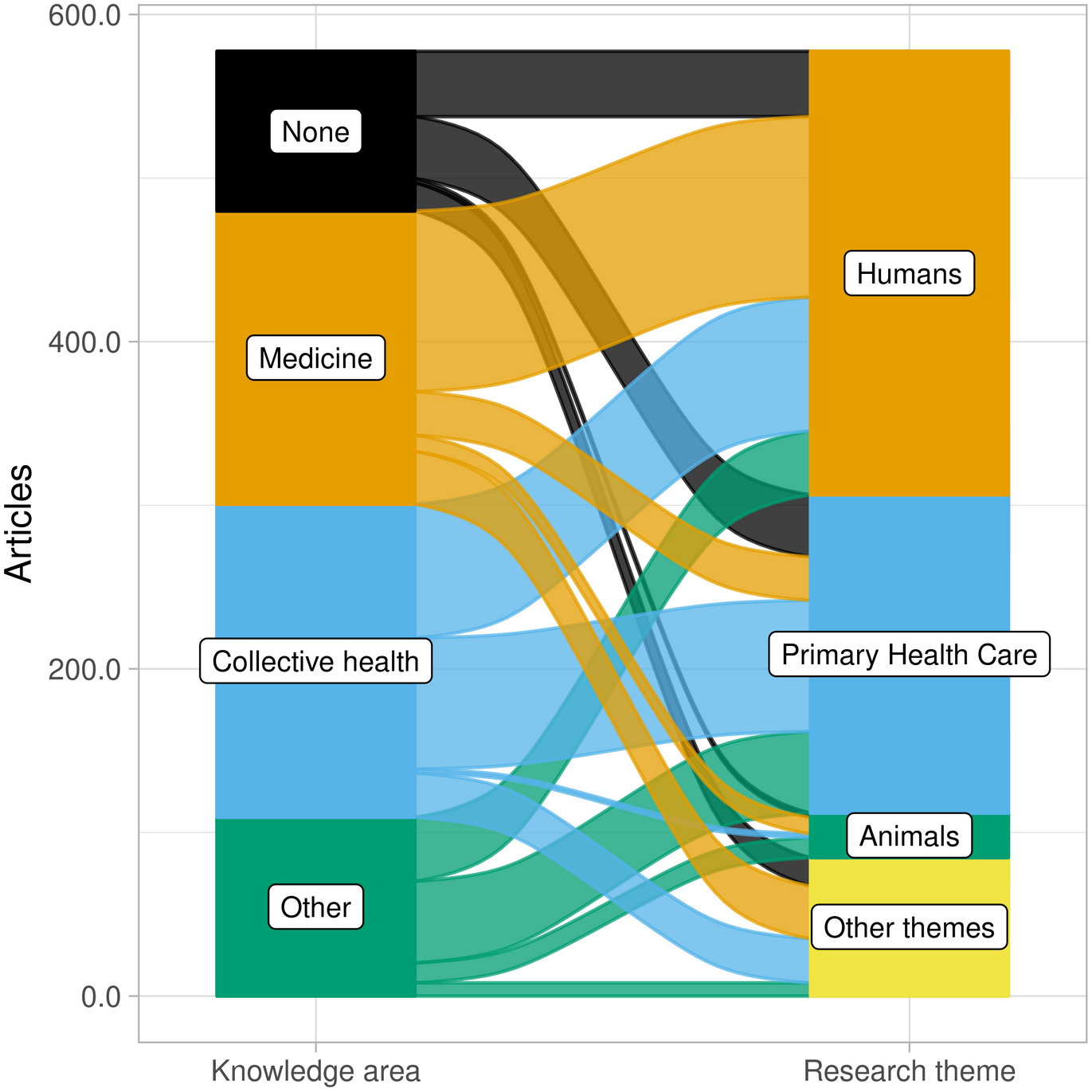
Alluvial plot of the weighted number of articles published by family and community physicians in Brazil, by knowledge area of the postgraduate degree and research theme of the article

Looking at the same numbers in another way, family and community physicians with a *stricto sensu* postgraduate degree in medicine usually authored articles in the *humans* research theme (110.4 out of 179.2). Meanwhile, the humans and the primary health care themes received equal attention from those with a degree in collective health (81.7 and 80.2, out of 191.3) or no degree (39.9 and 37.9, out of 97.7).

With regard to the level of the postgraduate degree, family and community physicians with a PhD accounted for most of the articles overall (51.2%) and in research themes *humans* (57.4%) and *animals* (78.4%) (Suppl. Table 2). In the *primary health care* theme, physicians with a PhD accounted for a similar proportion (39.1%) as those with a master’s (41.4%). Authors with a PhD were twice as likely to publish in the *humans* research theme as in the *primary health care* one, while those with a master’s or no postgraduate degree published equally in both themes.

Looking at these numbers in another way, family and community physicians with a PhD were twice as likely to publish articles in the *humans* research theme (155.6 out of 296.0) as in the *primary health care* (76.1). Meanwhile, those with a master’s published equally in the *humans* (75.6 articles out of 183.9) and *primary health care* (80.6) themes, as did those with no *stricto sensu* postgraduate degree.

## Discussion

### Summary of findings

Among family and community physicians in Brazil, the most common research themes were human health (*humans*) and primary care (*primary health care*), followed by preclinical studies in non-human animals (*animals*). Keywords on health conditions clustered mostly in the *humans* and *animals* themes, while keywords on health services, health systems and medical education clustered in the *primary health care* theme. These three main themes were those more connected to other themes through keyword co-occurrence; and *humans* and *primary health care* were the most developed ones, in terms of internal density of keyword co-occurrence. Over the years, *primary health care* saw the largest increase in size, and perhaps also in centrality and development. Looking at the author’s postgraduate trajectories, articles in the *primary health care* theme were much more likely to be authored by family and community physicians with a master’s or PhD in collective health.

### Strengths and weaknesses

Some methodological aspects should be considered when interpreting the research themes. One aspect is our choice of data sources. The Lattes Platform is a well established data source for studying the scholarly output of Brazilian researchers (Lane 2010; Leite et al. 2011; Mena-Chalco et al. 2014; Sidone et al. 2016), and active researchers can be expected to keep their publications list up to date. Starting from a national list of (potential) authors, instead of university departments or selected journals, allowed us to examine what are family and community physicians actually studying, and to correlate their output with their postgraduate trajectories. On the other hand, our data lacks any primary care research not done by Brazilian family and community physicians.

Obtaining our metadata from MEDLINE and, most importantly, LILACS allowed the inclusion of articles from all 14 journals publishing most of the articles by family and community physicians in Brazil (Fontenelle, Oliveira, et al. 2021). (One journal, *Revista AMRIGS*, stopped being index by LILACS in 2017.) In MEDLINE and LILACS both, librarians index journal articles following essentially the same methodology. Unfortunately, this approach made it impossible for us to separate original articles from editorials, opinion pieces and other non-research article types. Thus, our results relate more closely to scholarly output as a whole than to only the research output.

Another methodological aspect is the choice of clustering algorithm. For example, the size of research themes varied widely in our study but was homogeneous in Callon et al. (1991) and in Cobo et al. (2011). Furthermore, different research themes could have been derived from the same data by using different algorithms. In previous iterations of our methods, the *primary health care* theme remained consistent but the *humans* and *animals* themes varied in size and coherence.

Analyzing the data as a bipartite network, instead of concentrating on the keyword co-occurrence, allowed us to correlate the research themes with the postgraduate trajectories, and generally discarded less information even about the research themes themselves. Our choice of the dual-projection algorithm was heavily influenced by Arthur (2020), who concluded that applying the Louvain algorithm to the bipartite network was a reasonable but inferior alternative to applying it to both projections and then combining the result with hierarchical clustering (ie, the dual-projection algorithm). At the same time, we chose the Leiden algorithm over the Louvain one because of the advantages described by Traag et al. (2019).

Our choice of algorithm was not without disadvantages, though. Besides the necessity of custom code, the implemented approach is computationally intensive enough to be impractical outside research projects, and applying the algorithm separately for each time period would require finely tuning the resolution parameter again and again. Meanwhile, the leidenalg package for Python can directly apply the Leiden algorithm to bipartite networks without user-written functions, in a fraction of the time, and possibly using the overall resolution parameter for the individual time periods. We hope methodological researchers will compare the dual-projection to the direct Leiden algorithm for bipartite networks in the context of research themes, so that perhaps future research similar to ours can be more easily done.

Finally, another methodological aspect is the choice of the resolution parameter. Using a different value would result in a different number of research themes, splitting the *humans* theme in multiple sub-themes or merging it with *primary health care*. The interesting feature is not the number or size of the themes, but where lies the division between them.

### Interpretation

The *humans* research theme includes clinical research but also studies with a more epidemiological perspective. The difference between clinical and epidemiological research is often fuzzy: clinicians will do epidemiological research with clinical implications in mind, and most of the country’s primary care is community-based, so that practice-based research can be nearly population-based. This fuzziness might explain why *humans* is the largest research theme even though family and community physicians publish mostly in collective health journals (Fontenelle, Oliveira, et al. 2021).

It is telling that clinical research was found to be closer to epidemiological than to primary care research. The *primary health care* theme had no health conditions among its top keywords (at least up to two thirds of theme), as depicted in Suppl. Fig 2. This suggests family and community physicians in Brazil publish research on management, policy and medical education in primary care, as well as primary care as a concept, but little of their clinical research is specific to primary care. The lower average number of unique keywords per article in the *primary health care* theme seems to reflect this lack of clinical research in primary care.

The correlation between postgraduate trajectories and research themes deserves closer examination. Firstly, while only 2.7% of family and community physicians in Brazil hold a PhD degree (Fontenelle et al. 2020), they accounted for half the published articles. This finding emphasizes the role of *stricto sensu* postgraduate education (master’s and PhD courses) for research capacity in Brazil, and the possible consequences of family and community medicine or primary health care not being recognized as a knowledge area. All of the few postgraduate programs dedicated to primary health care are inside the “collective health” knowledge area, which might explain the disconnect between primary care and health conditions in the research themes. Furthermore, these programs conferred only master’s degrees until now, and their purpose is to qualify health professionals more for the health system than for academia (that is, they are professional *stricto sensu* postgraduate programs).

Secondly, while most family and community physicians in Brazil earn their *stricto sensu* postgraduate degrees in collective health (Fontenelle et al. 2020), those with a degree in medicine authored just as many articles. This is (at least in part) because medicine is almost as common as collective health for PhD degrees, and master’s degrees in collective health are less likely than those in medicine to be directed towards academia (Fontenelle et al. 2020).

Thirdly, family and community physicians with a *stricto sensu* postgraduate degree in collective health publish in the same themes as those with no postgraduate degree. This is in contrast with those with a degree in medicine publishing relatively more in the *humans* and less in the *primary health care* theme. This suggests earning a master’s or PhD degree in collective health allows these authors to keep their previous research interests, while earning a degree in medicine allows them to explore new research interests. Wenceslau et al. (2020) have proposed a decentralized professional master’s program in family and community medicine, as a specialty within medicine. If the proposal moves forward, it will be interesting to examine how the research themes of its alumni will compare to those of alumni of other postgraduate programs.

Fourthly, it doesn’t seem to matter whether the physicians specialized through medical residency or through certification (based on clinical experience, curriculum, and a test). Specialization through certification was just as common among family and community physicians as whole (Fontenelle et al. 2020) as among article authors. Although authors with certification were somewhat more common in the *primary health care* research theme, this can be explained by certification being available only since 2004, which happens to be when the national specialty journal (*Revista Brasileira de Medicina de Família e Comunidade* – RBMFC) was first published. In 2004–2008, family and community physicians in Brazil started publishing mostly in primary care journals (Fontenelle, Oliveira, et al. 2021), and published in the *primary health care* research theme thirteen times as much as in 1999–2003.

Finally, while most family and community physicians in Brazil are women (57%) (Fontenelle et al. 2020), most articles were published by men (64%). This authorship gap is even larger than the gap found in the top 3 family and community medicine journals from the United States (Jabbarpour et al. 2020). One explanation is that men have been more likely than women to obtain a master’s (adjusted odds ratio, 1.24) or PhD (1.86) degree (Fontenelle et al. 2020). Furthermore, whatever is holding women from pursuing *stricto sensu* postgraduate degrees might be holding them from performing research and publishing articles. As discussed in Fontenelle et al. (2020), this phenomenon seems to be more acute among family and community physicians than in the Brazilian society as a whole, and should be better understood.

Throughout the study period, there was not only an increase in the overall volume, as is often the case, but also changes in the research themes’ share of the scholarly output. There was a conspicuous increase in the volume of the primary health care research theme when SBMFC launched its journal, RBMFC, underlining the role of national specialty journals for strengthening research capacity. Contrary to the situation depicted by Harzheim et al. (2005), RBMFC is currently indexed in LILACS, another step for academic family medicine in Brazil. Unfortunately, RBMFC is not yet indexed in MEDLINE, a problem it shares with other family and community journals publishing non-English content (Mendis and Solangaarachchi 2005).

### Policy implications

The relative disconnect between primary care and clinical research in the scholarly output of family and community physicians suggests stakeholders need to promote the conduct of clinical research in the primary care setting. One way of doing so would be having primary care as a research line in medical *stricto sensu* postgraduate programs, or even having one or more programs dedicated to family and community medicine, as proposed by Wenceslau et al. (2020). However, the viability of such research lines or postgraduate programs might depend on changing how the government evaluates such programs. Currently, the evaluation of *stricto sensu* postgraduate programs depends heavily on citation metrics of the journals were the programs’ students and supervisors publish their research. Primary care research is usually more applied than the average biomedical research, resulting in a citation disadvantage (Donner and Schmoch 2020; Peleg and Shvartzman 2006) which might reflect negatively in the evaluation of medical postgraduate programs dedicated to primary care.

Another way of promoting clinical research in primary care would be taking steps to secure adequate funding. Primary care research funding is problematic worldwide, with primary care research being worse-funded than other clinical research (Goodyear-Smith and Mash 2016). The problem is made worse when there’s no funding line dedicated to primary care, as in the US National Institutes of Health (NIH): even though the NIH is the main source of research funding for university departments of family medicine in the US (Weidner et al. 2019), less than 1% of the projects funded by the NIH are primary care research (Westfall et al. 2021). In the Brazilian context, recognizing primary care or family and community medicine as a knowledge area would enable research funding agencies to dedicate part of their grants to clinical research in primary care.

Finally, another way would be fostering the creation of practice-based research networks (PBRNs) (L. W. Green 2008; Huas et al. 2019). Such networks allow academia to recruit patients and primary care practices for their research, and allow primary care practices to participate in the prioritization of research projects for real-world impact (Mold and Peterson 2005). While PBRNs have sometimes relied on volunteerism, the buy-in from societies and funding agencies is important for consolidating and scaling up (L. A. Green and Hickner 2006). In example, hiring practice facilitators is valuable for binding a PBRN together (Nagykaldi et al. 2006), as primary care providers and academics are often overwhelmed by clinical and teaching responsibilities (Brocato and Mavis 2005; Goodyear-Smith et al. 2016). Many of the good practices proposed for PBRNs (Dolor et al. 2015), also depend on some level of funding.

In the Brazilian context, the fragmentation of primary care management might be an obstacle to the implementation of PBRNs. Academia might have to negotiate with centralized provision programs such as *Mais Médicos* (More Doctors) and *Médicos pelo Brasil* (Doctors for Brazil), as well as its state-level equivalents, if large-scale PRBNs are to be feasible.

## Data Availability

The anonymized dataset is openly available in Zenodo. The underlying datasets are available upon reasonable request.

https://zenodo.org/record/5798132

## Declarations

### Availability of data

The anonymized dataset is openly available in Zenodo (https://zenodo.org/record/5798132), and the original ones are available upon reasonable request, as specified in the corresponding Zenodo deposits (https://doi.org/10.5281/zenodo.3376310 and https://doi.org/10.5281/zenodo.5797816).

### Authors’ contributions

Conceptualization: LFF, DJB, TDS. Data curation: LFF, SVR, MHMO. Formal analysis: LFF. Funding acquisition: LFF, MHMO. Investigation: LFF, SVR, MHMO. Methodology: LFF. Software: LFF. Visualization: LFF. Writing – original draft: LFF. Writing – review & edit: SVR, MHMO. DJB, TDS.

### Ethics approval

Research ethics committee of Universidade Vila Velha, report n° 3.033.911.

## Acknowledgments

MHMO was supported by a scientific initiation scholarship from Universidade Vila Velha.

## Footnotes

1 Cobo et al. (2011) refer to this density index as “Callon’s density”, citing Callon et al. (1991). However, the density index as described in Callon et al. (1991) matches more closely the usual network density index: summing the edge weights and dividing by the number of possible edges: k × (k-1), where k is the number of keywords. On the other hand, the dual-projection algorithm can yield arbitrarily large clusters, and articles can’t have an arbitrarily large number of keywords (say, many tens or hundreds). This would introduce a negative correlation between size and density of the research themes. Hence, we opted for density index put forward by Cobo et al. (2011).

## References

Almeida, M., Gusso, G., & Trindade, T. (2016). Primary care research in Brazil. In F. Goodyear-Smith & B. Mash (Eds.), International Perspectives on Primary Care Research (pp. 161–163). Boca Raton: CRC Press. Accessed 5 April 2020

Arthur, R. (2020). Modularity and projection of bipartite networks. Physica A: Statistical Mechanics and its Applications, 549, 124341. https://doi.org/10.1016/j.physa.2020.124341

Arya, N., Gibson, C., Ponka, D., Haq, C., Hansel, S., Dahlman, B., & Rouleau, K. (2017). Family medicine around the world: overview by region. Canadian Family Physician, 63(6), 436–441.

Barata, R. B., Aragão, E., Sousa, L.E.P.F.de, Santana, T. M., & Barreto, M. L. (2014). The configuration of the Brazilian scientific field. Anais Da Academia Brasileira De Ciencias, 86(1), 505–521. https://doi.org/10.1590/0001-3765201420130023

Barrat, A., Barthélemy, M., Pastor-Satorras, R., & Vespignani, A. (2004). The architecture of complex weighted networks. Proceedings of the National Academy of Sciences, 101(11), 3747–3752. https://doi.org/10.1073/pnas.0400087101

Beasley, J. W., Dovey, S., Geffen, L. N., Gómez-Clavelina, F. J., Haq, C. L., Inem, V., et al. (2004). The contribution of family doctors to primary care research: a global perspective from the International Federation of Primary Care Research Networks (IFPCRN). Primary Health Care Research & Development, 5(4), 307–316. https://doi.org/10.1191/1463423604pc221oa

Beasley, J. W., Starfield, B., van Weel, C., Rosser, W. W., & Haq, C. L. (2007). Global health and primary care research. Journal of the American Board of Family Medicine: JABFM, 20(6), 518–526. https://doi.org/10.3122/jabfm.2007.06.070172

Blondel, V. D., Guillaume, J.-L., Lambiotte, R., & Lefebvre, E. (2008). Fast unfolding of communities in large networks. Journal of Statistical Mechanics: Theory and Experiment, 2008(10), P10008. https://doi.org/10.1088/1742-5468/2008/10/P10008

Bonet, O. (2014). Os médicos da pessoa: um olhar antropológico sobre a medicina de família no Brasil e na Argentina. Rio de Janeiro: 7Letras. Accessed 11 August 2019

Brocato, J. J., & Mavis, B. (2005). The Research Productivity of Faculty in Family Medicine Departments at U.S. Medical Schools: A National Study. Academic Medicine, 80(3), 244–252.

Callon, M., Courtial, J. P., & Laville, F. (1991). Co-word analysis as a tool for describing the network of interactions between basic and technological research: The case of polymer chemsitry. Scientometrics, 22(1), 155–205. https://doi.org/10.1007/BF02019280

Ceitlin, J. (2006). La medicina familiar en América Latina. Presentación. Atención Primaria, 38(9), 511–514. https://doi.org/10.1157/13095056

Cobo, M. J., López-Herrera, A. G., Herrera-Viedma, E., & Herrera, F. (2011). An approach for detecting, quantifying, and visualizing the evolution of a research field: A practical application to the Fuzzy Sets Theory field. Journal of Informetrics, 5(1), 146–166. https://doi.org/10.1016/j.joi.2010.10.002

Csardi, G., & Nepusz, T. (2006). The igraph software package for complex network research. InterJournal, Complex Systems, 1695.

Dolor, R. J., Campbell-Voytal, K., Daly, J., Nagykaldi, Z. J., O’Beirne, M., Sterling, P., et al. (2015). Practice-based Research Network Research Good Practices (PRGPs): Summary of Recommendations. Clinical and Translational Science, 8(6), 638–646. https://doi.org/10.1111/cts.12317

Domínguez, N.M.S.de, Ponzo, J., Aranda, J. M. R., Haro, C. A. A., Ríos, M. E. R., Villarreal, P. V., et al. (2016). Research in Family and Community Medicine in Ibero-America. Revista Brasileira de Medicina de Família e Comunidade, 11(0), 64–74.

Donner, P., & Schmoch, U. (2020). The implicit preference of bibliometrics for basic research. Scientometrics, 124(2), 1411–1419. https://doi.org/10.1007/s11192-020-03516-3

Eck, N. J. van, & Waltman, L. (2009). How to normalize cooccurrence data? An analysis of some well-known similarity measures. Journal of the American Society for Information Science and Technology, 60(8), 1635–1651. https://doi.org/10.1002/asi.21075

Escobar, M. A. R., Rivera, L. M. M., Martínez, O. F., & Ortega, M.Á.F. (2018). Research applied to the territory from Family Medicine. Revista Brasileira de Medicina de Família e Comunidade, 13(1), 29–42. https://doi.org/10.5712/rbmfc13(1)1851

Everett, M. G., & Borgatti, S. P. (2013). The dual-projection approach for two-mode networks. Social Networks, 35(2), 204–210. https://doi.org/10.1016/j.socnet.2012.05.004

Falk, J. W. (2004). A Medicina de Família e Comunidade e sua entidade nacional: histórico e perspectivas. Revista Brasileira de Medicina de Família e Comunidade, 1(1), 5–10. https://doi.org/10.5712/rbmfc1(1)2

Fontenelle, L. F. (2021). Analytic code for “Research themes of family and community physicians in Brazil.” Zenodo. https://doi.org/10.5281/zenodo.5798092

Fontenelle, L. F., Oliveira, M. H. M. de, Rossi, S. V., Brandão, D. J., & Sarti, T. D. (2021). In which journals do family and community physicians in Brazil publish? the Trajetórias MFC project. Revista Brasileira de Medicina de Família e Comunidade, 16(43), 2589. https://doi.org/10.5712/rbmfc16(43)2589

Fontenelle, L. F., Rossi, S. V., & Oliveira, M.H.M.de. (2019). Postgraduate education among family and community physicians in Brazil: the Trajetórias MFC project. Zenodo. https://doi.org/10.5281/zenodo.3376310

Fontenelle, L. F., Rossi, S. V., & Oliveira, M.H.M. de. (2021a, December 22). Journal articles published by family and community physicians from Brazil up to 2018. Zenodo. https://doi.org/10.5281/ZENODO.5797816

Fontenelle, L. F., Rossi, S. V., Oliveira, M.H.M. de, Brandão, D. J., & Sarti, T. D. (2020). Postgraduate education among family and community physicians in Brazil: the Trajetórias MFC project. Family Medicine and Community Health, 8(3), e000321. https://doi.org/10.1136/fmch-2020-000321

Fontenelle, L. F., Rossi, S. V., & Oliveira, M. H. M. (2021b, December 22). Anonymized dataset for “Research themes of family and community physicians in Brazil.” Zenodo. https://doi.org/10.5281/zenodo.5798132

Freeman, T. (2012). Family Medicine’s academic contributions. Family Medicine Research Days, İzmir, Turkey. Türkiye aile hekimliği dergisi, 16(4), 181–198. https://doi.org/10.2399/tahd.12.181

Freeman, T., & McWhinney, I. R. (2016). McWhinney’s textbook of family medicine (4th edition.). Oxford ; New York: Oxford University Press.

Fruchterman, T. M. J., & Reingold, E. M. (1991). Graph drawing by force-directed placement. Software: Practice and Experience, 21(11), 1129–1164. https://doi.org/10.1002/spe.4380211102

Glanville, J., Kendrick, T., McNally, R., Campbell, J., & Hobbs, F. R. (2011). Research output on primary care in Australia, Canada, Germany, the Netherlands, the United Kingdom, and the United States: bibliometric analysis. BMJ, 342, d1028. https://doi.org/10.1136/bmj.d1028

Glänzel, W., Leta, J., & Thijs, B. (2006). Science in Brazil. Part 1: A macro-level comparative study. Scientometrics, 67(1), 67–86. https://doi.org/10.1007/s11192-006-0055-7

Goodyear-Smith, F., & Mash, B. (2016). Introduction. In F. Goodyear-Smith & B. Mash (Eds.), International Perspectives on Primary Care Research (pp. 161–163). Boca Raton: CRC Press. Accessed 5 April 2020

Goodyear-Smith, F., Mash, B., & Mash, B. (Eds.). (2016). Who conducts primary care research? In International Perspectives on Primary Care Research (pp. 11–18). CRC Press. Accessed 5 April 2020

Grácio, M. C. C., & Oliveira, E.F.T.de. (2014). Indicadores cientométricos normalizados: um estudo na produção científica brasileira internacional (1996 a 2011). Perspectivas em Ciência da Informação, 19(3), 118–133. https://doi.org/10.1590/1981-5344/1898

Green, L. A., & Hickner, J. (2006). A Short History of Primary Care Practice-based Research Networks: From Concept to Essential Research Laboratories. The Journal of the American Board of Family Medicine, 19(1), 1–10. https://doi.org/10.3122/jabfm.19.1.1

Green, L. W. (2008). Making research relevant: if it is an evidence-based practice, where’s the practice-based evidence? Family Practice, 25(suppl_1), i20–i24. https://doi.org/10.1093/fampra/cmn055

Gupta, A., Gray, C. S., Landes, M., Sridharan, S., & Bhattacharyya, O. (2021). Family medicine: An evolving field around the world. Canadian Family Physician, 67(9), 647–651. https://doi.org/10.46747/cfp.6709647

Hagen, N. T. (2008). Harmonic Allocation of Authorship Credit: Source-Level Correction of Bibliometric Bias Assures Accurate Publication and Citation Analysis. PLOS ONE, 3(12), e4021. https://doi.org/10.1371/journal.pone.0004021

Hagen, N. T. (2010). Harmonic publication and citation counting: sharing authorship credit equitably – not equally, geometrically or arithmetically. Scientometrics, 84(3), 785–793. https://doi.org/10.1007/s11192-009-0129-4

Hagen, N. T. (2013). Harmonic coauthor credit: A parsimonious quantification of the byline hierarchy. Journal of Informetrics, 7(4), 784–791. https://doi.org/10.1016/j.joi.2013.06.005

Harzheim, E., Stein, A. T., Álvarez Dardet, C., Cantero, M. T. R., Kruse, C. K., Vidal, T. B., & Nava, T. R. (2005). Revisão sistemática sobre aspectos metodológicos das pesquisas em atenção primária no Brasil. Revista dad AMRIGS, 49(4), 248–252.

Huas, C., Petek, D., Diaz, E., Muñoz-Perez, M. A., Torzsa, P., & Collins, C. (2019). Strategies to improve research capacity across European general practice: The views of members of EGPRN and Wonca Europe. European Journal of General Practice, 0(0), 1–7. https://doi.org/10.1080/13814788.2018.1546282

Jabbarpour, Y., Wilkinson, E., Coffman, M., & Mieses, A. (2020). Has Female Authorship in Family Medicine Research Evolved Over Time? Annals of Family Medicine, 18(6), 496–502. https://doi.org/10.1370/afm.2584

Lane, J. (2010). Let’s make science metrics more scientific. Nature, 464, 488–489. https://doi.org/10.1038/464488a

Leite, P., Mugnaini, R., & Leta, J. (2011). A new indicator for international visibility: exploring Brazilian scientific community. Scientometrics, 88(1), 311. https://doi.org/10.1007/s11192-011-0379-9

Leta, J., Thijs, B., & Glänzel, W. (2013). A macro-level study of science in Brazil: seven years later. Encontros Bibli, 18(36), 51–66. https://doi.org/10.5007/1518-2924.2013v18n36p51

López-Torres Hidalgo, J., Basora Gallisà, J., Orozco Beltrán, D., & Bellón Saameño, J.Á. (2014). Mapa bibliométrico de la investigación realizada en atención primaria en España durante el periodo 2008-2012. Atención Primaria, 46(10), 541–548. https://doi.org/10.1016/j.aprim.2014.02.007

López-Torres Hidalgo, J., Párraga Martínez, I., Martín Álvarez, R., & Tranche Iparraguirre, S. (2020). Mapa bibliométrico de la investigación realizada en atención primaria en España durante el periodo 2013-2017. Atención Primaria, 52(7), 469–476. https://doi.org/10.1016/j.aprim.2019.08.002

MacAuley, D. (2011). Brazilian Family Medicine and academic excellence. Revista Brasileira de Medicina de Família e Comunidade, 6(20), 171–174. https://doi.org/10.5712/rbmfc6(20)403

Mash, R., Almeida, M., Wong, W. C. W., Kumar, R., & von Pressentin, K. B. (2015). The roles and training of primary care doctors: China, India, Brazil and South Africa. Human Resources for Health, 13(1), 93. https://doi.org/10.1186/s12960-015-0090-7

McWhinney, I. R. (1996). The importance of being different. The British Journal of General Practice, 46(408), 433–436.

McWhinney, I. R. (2001a). Why are we doing so little clinical research? Part 1: Clinical descriptive research. Canadian Family Physician, 47, 1701–1715.

McWhinney, I. R. (2001b). Why are we doing so little clinical research? Part 2: Why clinical research is neglected. Canadian Family Physician Medecin De Famille Canadien, 47, 1944–1946, 1952–1955.

Melamed, D. (2014). Community Structures in Bipartite Networks: A Dual-Projection Approach. PLOS ONE, 9(5), e97823. https://doi.org/10.1371/journal.pone.0097823

Mena-Chalco, J. P., Digiampietri, L. A., Lopes, F. M., & Cesar, R. M. (2014). Brazilian Bibliometric Coauthorship Networks. Journal of the Association for Information Science and Technology, 65(7), 1424–1445. https://doi.org/10.1002/asi.23010

Mendis, K., & Solangaarachchi, I. (2005). PubMed perspective of family medicine research: where does it stand? Family Practice, 22(5), 570–575. https://doi.org/10.1093/fampra/cmi085

Mold, J. W., & Peterson, K. A. (2005). Primary Care Practice-Based Research Networks: Working at the Interface Between Research and Quality Improvement. The Annals of Family Medicine, 3(suppl 1), S12–S20. https://doi.org/10.1370/afm.303

Muldoon, L. K., Hogg, W. E., & Levitt, M. (2006). Primary care (PC) and primary health care (PHC). What is the difference? Canadian Journal of Public Health = Revue Canadienne De Sante Publique, 97(5), 409–411.

Nagykaldi, Z., Mold, J. W., Robinson, A., Niebauer, L., & Ford, A. (2006). Practice Facilitators and Practice-based Research Networks. The Journal of the American Board of Family Medicine, 19(5), 506–510. https://doi.org/10.3122/jabfm.19.5.506

Newman, M. E. J. (2004). Analysis of weighted networks. Physical Review E, 70(5), 056131. https://doi.org/10.1103/PhysRevE.70.056131

Newman, M. E. J., & Girvan, M. (2004). Finding and evaluating community structure in networks. Physical Review E, 69(2), 026113. https://doi.org/10.1103/PhysRevE.69.026113

Okabe, M., & Ito, K. (2002). How to make figures and presentations that are friendly to color blind people. University of Tokyo. https://jfly.uni-koeln.de/html/color_blind/

Ortega, M.Á.F., Velazco, G. R., Coria, A. E. I., & Prato, J. B. R. (2016). Producción y difusión de conocimientos en medicina familiar en Iberoamérica. Revista Brasileira de Medicina de Família e Comunidade, 11, 71–87. https://doi.org/10.5712/rbmfc11(0)1280

Osmo, A., & Schraiber, L. B. (2015). The field of Collective Health: definitions and debates on its constitution. Saúde e Sociedade, 24, 205–218. https://doi.org/10.1590/S0104-12902015S01018

Peleg, R., & Shvartzman, P. (2006). Where Should Family Medicine Papers be Published— Following the Impact Factor? The Journal of the American Board of Family Medicine, 19(6), 633–636. https://doi.org/10.3122/jabfm.19.6.633

Pires-Alves, F. A., & Paiva, C. H. A. (2021). Entre a ausência em Alma-Ata e o Prevsaúde:a atenção primária à saúde no ocaso da ditadura. História, Ciências, Saúde-Manguinhos, 28(3), 643–659. https://doi.org/10.1590/s0104-59702021000300002

Ponka, D., Rouleau, K., Arya, N., Redwood-Campbell, L., Woollard, R., Siedlecki, B., & Dunikowski, L. (2015). Developing the evidentiary basis for family medicine in the global context: The Besrour Papers: a series on the state of family medicine in the world. Canadian Family Physician Medecin De Famille Canadien, 61(7), 596–600.

Pons, P., & Latapy, M. (2006). Computing Communities in Large Networks Using Random Walks. Journal of Graph Algorithms and Applications, 10(2), 191–218. https://doi.org/10.7155/jgaa.00124

Rodrigues, R. D. (2007). Programa de Residência em Medicina de Família e Comunidade da UERJ: uma perspectiva histórica. Revista Brasileira de Medicina de Família e Comunidade, 3(11), 149–156. https://doi.org/10.5712/rbmfc3(11)333

Rogers, F. B. (1963). Medical subject headings. Bulletin of the Medical Library Association, 51(1), 114–116.

Romano, V. F. (2008). In search of identity for the family doctor. Physis: Revista de Saúde Coletiva, 18(1), 13–25. https://doi.org/10.1590/S0103-73312008000100002

Sidone, O. J. G., Haddad, E. A., & Mena-Chalco, J. P. (2016). A ciência nas regiões brasileiras: evolução da produção e das redes de colaboração científica. Transinformação, 28(1), 15–32. https://doi.org/10.1590/2318-08892016002800002

Siqueira, M. B. (2019). Sucupira - A Platform for the Evaluation of Graduate Education in Brazil. Procedia Computer Science, 146, 247–255. https://doi.org/10.1016/j.procs.2019.01.081

Sparks, B. L. W., & Gupta, S. K. (2004). Research in Family Medicine in Developing Countries. Annals of Family Medicine, 2(suppl 2), S55–S59. https://doi.org/10.1370/afm.192

Starfield, B., Shi, L., & Macinko, J. (2005). Contribution of primary care to health systems and health. The Milbank quarterly, 83(3), 457–502. https://doi.org/10.1111/j.1468-0009.2005.00409.x

Steijn, M. P. A. (2021). Improvement on the association strength: Implementing a probabilistic measure based on combinations without repetition. Quantitative Science Studies, 2(2), 778–794. https://doi.org/10.1162/qss_a_00122

Traag, V. A., Van Dooren, P., & Nesterov, Y. (2011). Narrow scope for resolution-limit-free community detection. Physical Review E, 84(1), 016114. https://doi.org/10.1103/PhysRevE.84.016114

Traag, V. A., Waltman, L., & van Eck, N. J. (2019). From Louvain to Leiden: guaranteeing well-connected communities. Scientific Reports, 9(1), 5233. https://doi.org/10.1038/s41598-019-41695-z

Trefethen, L. N., & Bau III, D. (1997). Numerical linear algebra. Siam.

van der Zee, J., Kroneman, M., & Bolíbar, B. (2003). Conditions for research in general practice. Can the Dutch and British experiences be applied to other countries, for example Spain? The European Journal of General Practice, 9(2), 41–47. https://doi.org/10.3109/13814780309160401

Vieira-da-Silva, L. M., & Pinell, P. (2014). The genesis of collective health in Brazil. Sociology of Health & Illness, 36(3), 432–446. https://doi.org/10.1111/1467-9566.12069

Weidner, A., Peterson, L. E., Mainous, A. G., Datta, A., & Ewigman, B. (2019). The Current State of Research Capacity in US Family Medicine Departments. Family Medicine, 51(2), 112–119. https://doi.org/10.22454/FamMed.2019.180310

Wenceslau, L. D., Sarti, T. D., & Trindade, T.G.da. (2020). Reflections and proposals for the establishment of Family and Community Medicine Master’s Programs in Brazil. Ciência & Saúde Coletiva, 25(4), 1281–1292. https://doi.org/10.1590/1413-81232020254.29802019

Westfall, J. M., Wittenberg, H. R., & Liaw, W. (2021). Time to Invest in Primary Care Research— Commentary on Findings from an Independent Congressionally Mandated Study. Journal of General Internal Medicine. https://doi.org/10.1007/s11606-020-06560-0

Wickham, H. (2016). ggplot2: Elegant Graphics for Data Analysis (2nd ed.). Cham: Springer International Publishing. https://doi.org/10.1007/978-3-319-24277-4

