## Supplementary material for "Research themes of family and community physicians in Brazil": Suppl

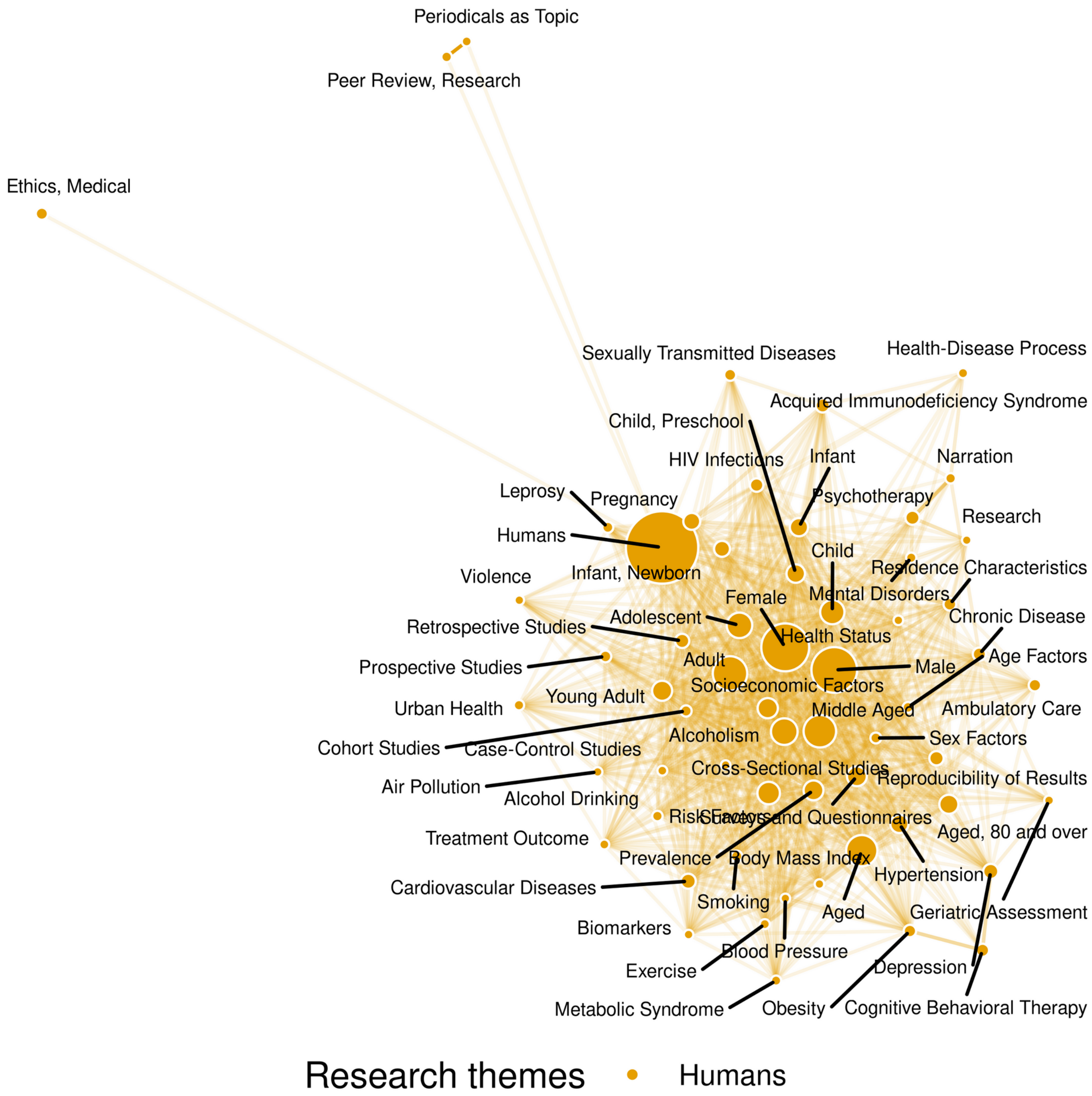


*Suppl. Figure 1 – Diagram of keyword co-occurrence for the most common keywords, amounting to four fifths of the "humans" theme. Bubble sizes and edges' opacity are proportional to weighted frequency.*

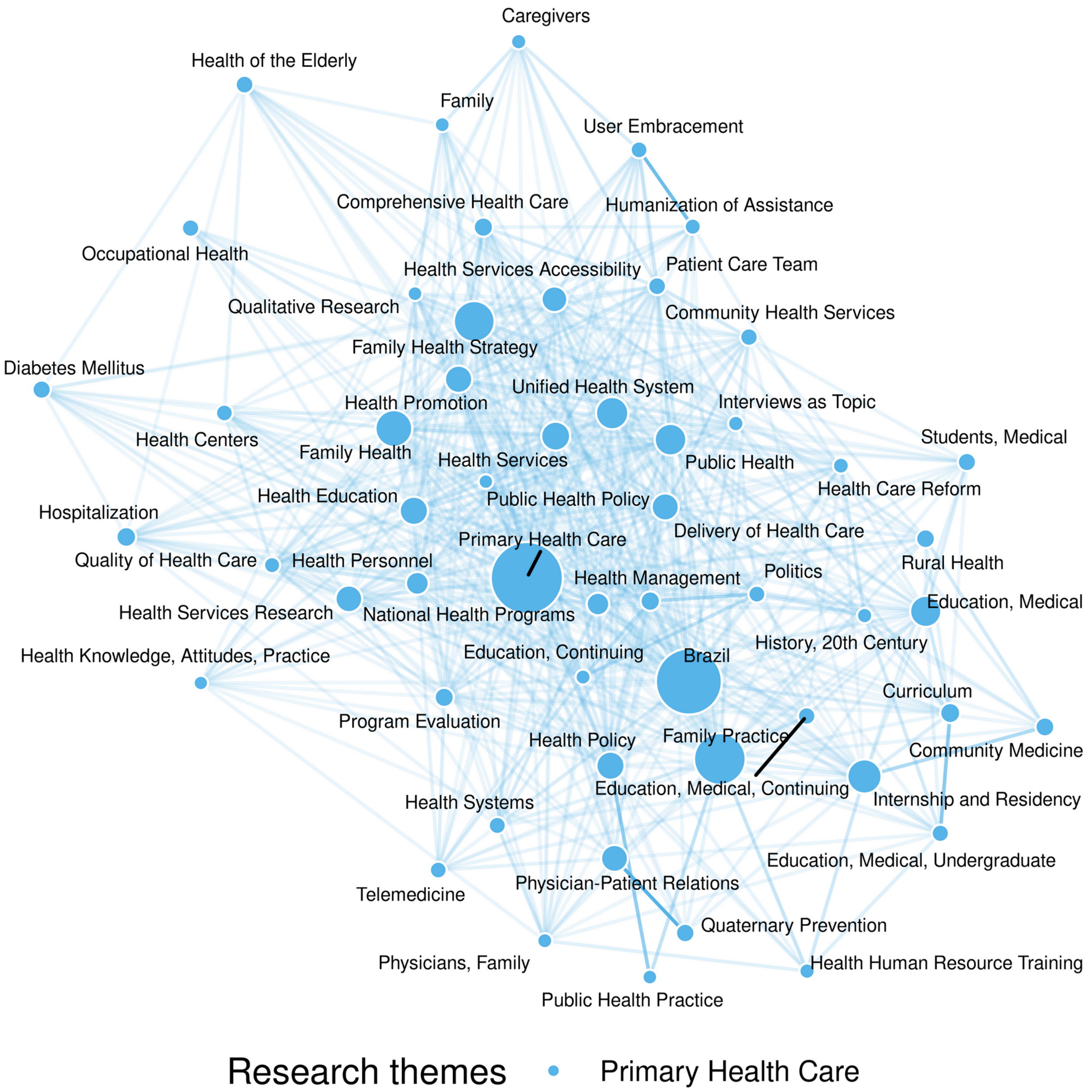


*Suppl. Figure 2 – Diagram of keyword co-occurrence for the most common keywords, amounting to two thirds in the "primary health care" theme. Bubble sizes and edges' opacity are proportional to weighted frequency.*

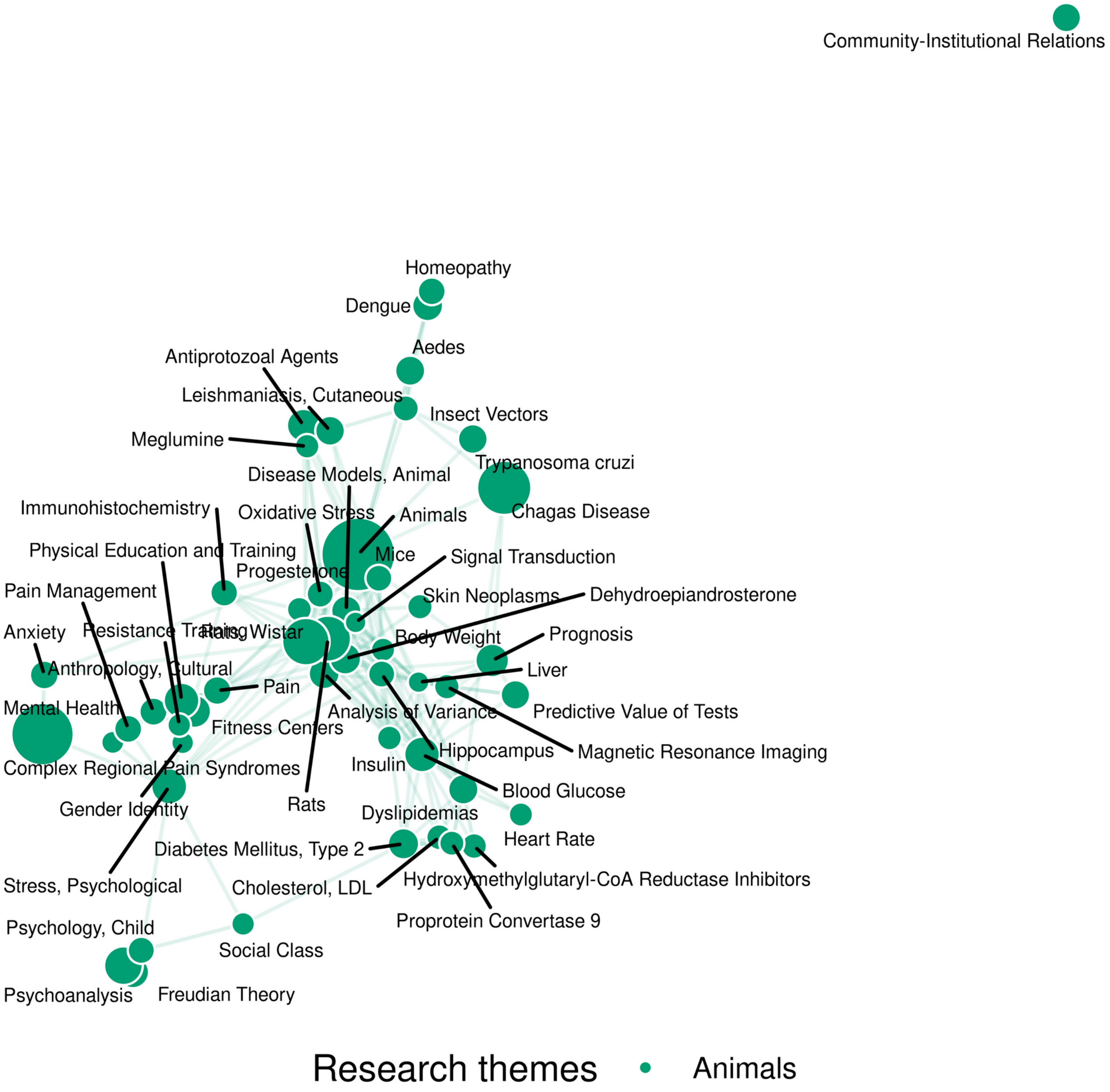


*Suppl. Figure 3 – Diagram of keyword co-occurrence for the most common keywords, amounting to half the "animals" theme. Bubble sizes and edges' opacity are proportional to weighted frequency.*

*Suppl. Table 1 – Characteristics of the research themes of family and community physicians in Brazil, up to 2018*

| ID | Name * | Articles unique | Articles weighted | Authorship weight (mean) | Keywords per article (mean) | Keywords unique | Keywords weighted | Centrality | Density |
|---|---|---|---|---|---|---|---|---|---|
| 1 | Humans | 1005 | 271.1 | 27.0% | 10.4 | 744 | 242.7 | 4776.3 | 36.4 |
| 2 | Primary Health Care | 419 | 194.5 | 46.4% | 5.5 | 586 | 226.0 | 2302.1 | 33.1 |
| 3 | Animals | 156 | 27.2 | 17.4% | 12.4 | 625 | 31.5 | 3329.3 | 12.4 |
| 4 | Malaria, Vivax | 34 | 4.0 | 11.7% | 15.9 | 120 | 4.5 | 992.9 | 9.4 |
| 5 | Sensitivity and Specificity | 17 | 5.0 | 29.4% | 10.1 | 41 | 3.2 | 365.5 | 8.2 |
| 6 | Severity of Illness Index | 14 | 2.1 | 15.2% | 10.5 | 69 | 4.4 | 560.2 | 7.5 |
| 7 | Influenza, Human | 13 | 3.4 | 26.3% | 8.6 | 16 | 1.4 | 134.9 | 13.5 |
| 8 | Educational Measurement | 12 | 3.3 | 27.6% | 10.2 | 9 | 1.0 | 30.4 | 15.2 |
| 9 | Zika Virus | 3 | 1.7 | 56.8% | 4.4 | 11 | 2.3 | 95.9 | 13.0 |
| 10 | Breast Feeding | 13 | 2.5 | 19.2% | 8.1 | 13 | 1.4 | 59.0 | 14.3 |
| 11 | Accidental Falls | 5 | 1.0 | 20.3% | 7.9 | 11 | 1.0 | 57.4 | 4.2 |
| 12 | Drug Utilization | 4 | 1.0 | 25.3% | 8.2 | 12 | 0.9 | 71.2 | 5.6 |
| 13 | Dementia | 0 | 0.0 | | | 17 | 1.7 | 54.2 | 0.0 |
| 14 | | 5 | 1.7 | 33.4% | 10.7 | 0 | 0.0 | 0.0 | 0.0 |
| 15 | Referral and Consultation | 2 | 1.1 | 52.8% | 5.2 | 17 | 0.5 | 59.6 | 2.9 |
| 16 | Practice Patterns, Physicians' | 0 | 0.0 | | | 12 | 1.5 | 49.0 | 0.0 |
| 17 | Evidence-Based Medicine | 0 | 0.0 | | | 11 | 1.5 | 81.5 | 0.0 |
| 18 | Community Health Workers | 0 | 0.0 | | | 10 | 1.5 | 63.4 | 0.0 |
| 19 | Episode of Care | 0 | 0.0 | | | 15 | 1.4 | 132.9 | 0.0 |
| 20 | Ethics, Clinical | 0 | 0.0 | | | 16 | 1.4 | 215.5 | 0.0 |
| 21 | Time Factors | 0 | 0.0 | | | 17 | 1.3 | 197.9 | 0.0 |

| ID | Name * | Articles unique | Articles weighted | Authorship weight (mean) | Keywords per article (mean) | Keywords unique | Keywords weighted | Centrality | Density |
|---|---|---|---|---|---|---|---|---|---|
| 22 | HIV | 0 | 0.0 | | | 9 | 1.3 | 36.4 | 0.0 |
| 23 | Quality of Life | 0 | 0.0 | | | 8 | 1.3 | 60.7 | 0.0 |
| 24 | | 4 | 1.3 | 32.3% | 5.2 | 0 | 0.0 | 0.0 | 0.0 |
| 25 | Aging | 0 | 0.0 | | | 8 | 1.3 | 31.5 | 0.0 |
| 26 | Dental Caries | 0 | 0.0 | | | 10 | 1.2 | 52.8 | 0.0 |
| 27 | Disease Outbreaks | 0 | 0.0 | | | 13 | 1.2 | 133.4 | 0.0 |
| 28 | Urban Population | 0 | 0.0 | | | 11 | 1.2 | 69.2 | 0.0 |
| 29 | Students | 0 | 0.0 | | | 9 | 1.2 | 56.0 | 0.0 |
| 30 | | 4 | 1.2 | 28.9% | 23.3 | 0 | 0.0 | 0.0 | 0.0 |
| 31 | | 3 | 1.1 | 37.6% | 7.7 | 0 | 0.0 | 0.0 | 0.0 |
| 32 | Frail Elderly | 0 | 0.0 | | | 6 | 1.1 | 45.5 | 0.0 |
| 33 | Developing Countries | 0 | 0.0 | | | 15 | 1.1 | 109.9 | 0.0 |
| 34 | | 5 | 1.0 | 20.8% | 14.5 | 0 | 0.0 | 0.0 | 0.0 |
| 35 | Practice Guidelines as Topic | 1 | 0.3 | 27.6% | 9.0 | 8 | 0.7 | 45.0 | 6.2 |
| 36 | | 1 | 1.0 | 100.0% | 7.0 | 0 | 0.0 | 0.0 | 0.0 |
| 37 | | 1 | 1.0 | 100.0% | 6.0 | 0 | 0.0 | 0.0 | 0.0 |
| 38 | | 1 | 1.0 | 100.0% | 13.0 | 0 | 0.0 | 0.0 | 0.0 |
| 39 | | 1 | 1.0 | 100.0% | 9.0 | 0 | 0.0 | 0.0 | 0.0 |
| 40 | Attitude of Health Personnel | 0 | 0.0 | | | 24 | 0.9 | 178.3 | 0.0 |
| 41 | Antiretroviral Therapy, Highly Active | 0 | 0.0 | | | 21 | 0.9 | 269.0 | 0.0 |
| 42 | Health Surveys | 0 | 0.0 | | | 11 | 0.9 | 99.7 | 0.0 |
| 43 | Trust | 3 | 0.5 | 17.5% | 16.2 | 16 | 0.4 | 165.4 | 3.0 |

| ID | Name * | Articles unique | Articles weighted | Authorship weight (mean) | Keywords per article (mean) | Keywords unique | Keywords weighted | Centrality | Density |
|---|---|---|---|---|---|---|---|---|---|
| 44 | Coinfection | 0 | 0.0 | | | 8 | 0.9 | 71.2 | 0.0 |
| 45 | Diet | 0 | 0.0 | | | 10 | 0.9 | 133.5 | 0.0 |
| 46 | Models, Statistical | 0 | 0.0 | | | 12 | 0.9 | 108.1 | 0.0 |
| 47 | Epidemiology, Descriptive | 1 | 0.4 | 36.0% | 8.0 | 6 | 0.5 | 50.7 | 5.4 |
| 48 | Infant Mortality | 0 | 0.0 | | | 9 | 0.9 | 54.5 | 0.0 |
| 49 | | 3 | 0.9 | 28.9% | 5.9 | 0 | 0.0 | 0.0 | 0.0 |
| 50 | | 2 | 0.9 | 43.0% | 9.4 | 0 | 0.0 | 0.0 | 0.0 |
| 51 | Automobile Driving | 0 | 0.0 | | | 18 | 0.8 | 144.2 | 0.0 |
| 52 | Patient Compliance | 0 | 0.0 | | | 18 | 0.8 | 178.3 | 0.0 |
| 53 | Liver Cirrhosis | 0 | 0.0 | | | 13 | 0.8 | 167.4 | 0.0 |
| 54 | | 1 | 0.8 | 81.8% | 8.0 | 0 | 0.0 | 0.0 | 0.0 |
| 55 | | 4 | 0.8 | 20.3% | 17.8 | 0 | 0.0 | 0.0 | 0.0 |
| 56 | Employee Performance Appraisal | 0 | 0.0 | | | 12 | 0.8 | 99.2 | 0.0 |
| 57 | Depression, Postpartum | 0 | 0.0 | | | 13 | 0.8 | 135.9 | 0.0 |
| 58 | Substance-Related Disorders | 0 | 0.0 | | | 8 | 0.8 | 73.6 | 0.0 |
| 59 | Blood Pressure Monitoring, Ambulatory | 0 | 0.0 | | | 12 | 0.8 | 136.7 | 0.0 |
| 60 | Incidence | 0 | 0.0 | | | 25 | 0.8 | 219.0 | 0.0 |
| 61 | Self Concept | 0 | 0.0 | | | 9 | 0.8 | 54.7 | 0.0 |
| 62 | | 1 | 0.8 | 76.7% | 12.0 | 0 | 0.0 | 0.0 | 0.0 |
| 63 | | 3 | 0.8 | 25.2% | 18.4 | 0 | 0.0 | 0.0 | 0.0 |
| 64 | Chagas Cardiomyopathy | 0 | 0.0 | | | 26 | 0.8 | 216.2 | 0.0 |
| 65 | Inflammation | 1 | 0.1 | 6.4% | 15.0 | 27 | 0.7 | 392.4 | 2.1 |

| ID | Name * | Articles unique | Articles weighted | Authorship weight (mean) | Keywords per article (mean) | Keywords unique | Keywords weighted | Centrality | Density |
|---|---|---|---|---|---|---|---|---|---|
| 66 | AIDS-Related Opportunistic Infections | 1 | 0.1 | 13.6% | 18.0 | 13 | 0.6 | 144.8 | 3.6 |
| 67 | Consensus | 0 | 0.0 | | | 12 | 0.7 | 124.9 | 0.0 |
| 68 | Epidemiologic Methods | 0 | 0.0 | | | 13 | 0.7 | 102.4 | 0.0 |
| 69 | | 2 | 0.7 | 34.4% | 12.5 | 0 | 0.0 | 0.0 | 0.0 |
| 70 | Translations | 0 | 0.0 | | | 16 | 0.7 | 130.5 | 0.0 |
| 71 | Cluster Analysis | 0 | 0.0 | | | 22 | 0.7 | 209.3 | 0.0 |
| 72 | Body Constitution | 1 | 0.5 | 50.0% | 6.0 | 2 | 0.2 | 0.5 | 11.1 |
| 73 | | 2 | 0.7 | 33.3% | 12.5 | 0 | 0.0 | 0.0 | 0.0 |
| 74 | | 2 | 0.7 | 33.0% | 8.0 | 0 | 0.0 | 0.0 | 0.0 |
| 75 | | 3 | 0.6 | 21.1% | 5.6 | 0 | 0.0 | 0.0 | 0.0 |
| 76 | Acute Kidney Injury | 1 | 0.1 | 10.2% | 14.0 | 25 | 0.5 | 249.9 | 2.5 |
| 77 | Antimalarials | 0 | 0.0 | | | 16 | 0.6 | 115.7 | 0.0 |
| 78 | | 4 | 0.6 | 15.2% | 14.5 | 0 | 0.0 | 0.0 | 0.0 |
| 79 | Child Abuse | 0 | 0.0 | | | 9 | 0.6 | 36.7 | 0.0 |
| 80 | House Calls | 1 | 0.4 | 36.0% | 9.0 | 2 | 0.2 | 7.6 | 7.4 |
| 81 | Tomography, X-Ray Computed | 0 | 0.0 | | | 13 | 0.6 | 103.8 | 0.0 |
| 82 | Parents | 0 | 0.0 | | | 6 | 0.6 | 54.0 | 0.0 |
| 83 | Acute Coronary Syndrome | 1 | 0.4 | 40.9% | 8.0 | 2 | 0.2 | 7.8 | 8.3 |
| 84 | | 1 | 0.6 | 56.2% | 11.0 | 0 | 0.0 | 0.0 | 0.0 |
| 85 | | 2 | 0.6 | 27.6% | 9.4 | 0 | 0.0 | 0.0 | 0.0 |
| 86 | Recurrence | 0 | 0.0 | | | 9 | 0.6 | 70.7 | 0.0 |
| 87 | | 1 | 0.5 | 54.4% | 14.0 | 0 | 0.0 | 0.0 | 0.0 |

| ID | Name * | Articles unique | Articles weighted | Authorship weight (mean) | Keywords per article (mean) | Keywords unique | Keywords weighted | Centrality | Density |
|---|---|---|---|---|---|---|---|---|---|
| 88 | Consumer Behavior | 0 | 0.0 | | | 10 | 0.5 | 84.4 | 0.0 |
| 89 | | 1 | 0.5 | 52.0% | 10.0 | 0 | 0.0 | 0.0 | 0.0 |
| 90 | Integrality in Health | 0 | 0.0 | | | 4 | 0.5 | 18.0 | 0.0 |
| 91 | Diabetes Complications | 0 | 0.0 | | | 12 | 0.5 | 49.7 | 0.0 |
| 92 | Bodily Secretions | 1 | 0.4 | 36.0% | 15.0 | 6 | 0.1 | 5.6 | 5.7 |
| 93 | Survival Rate | 0 | 0.0 | | | 12 | 0.5 | 84.3 | 0.0 |
| 94 | | 1 | 0.5 | 50.0% | 14.0 | 0 | 0.0 | 0.0 | 0.0 |
| 95 | | 1 | 0.5 | 50.0% | 7.0 | 0 | 0.0 | 0.0 | 0.0 |
| 96 | | 1 | 0.5 | 50.0% | 7.0 | 0 | 0.0 | 0.0 | 0.0 |
| 97 | | 1 | 0.5 | 50.0% | 8.0 | 0 | 0.0 | 0.0 | 0.0 |
| 98 | | 1 | 0.5 | 50.0% | 9.0 | 0 | 0.0 | 0.0 | 0.0 |
| 99 | | 1 | 0.5 | 50.0% | 13.0 | 0 | 0.0 | 0.0 | 0.0 |
| 100 | | 1 | 0.5 | 50.0% | 6.0 | 0 | 0.0 | 0.0 | 0.0 |
| 101 | | 1 | 0.5 | 50.0% | 13.0 | 0 | 0.0 | 0.0 | 0.0 |
| 102 | | 1 | 0.5 | 50.0% | 11.0 | 0 | 0.0 | 0.0 | 0.0 |
| 103 | | 1 | 0.5 | 50.0% | 11.0 | 0 | 0.0 | 0.0 | 0.0 |
| 104 | | 1 | 0.5 | 50.0% | 6.0 | 0 | 0.0 | 0.0 | 0.0 |
| 105 | | 1 | 0.5 | 50.0% | 11.0 | 0 | 0.0 | 0.0 | 0.0 |
| 106 | | 1 | 0.5 | 50.0% | 8.0 | 0 | 0.0 | 0.0 | 0.0 |
| 107 | Neurofibromatosis 1 | 0 | 0.0 | | | 21 | 0.5 | 165.1 | 0.0 |
| 108 | | 1 | 0.5 | 48.0% | 12.0 | 0 | 0.0 | 0.0 | 0.0 |
| 109 | Acute Disease | 0 | 0.0 | | | 13 | 0.5 | 122.1 | 0.0 |

| ID | Name * | Articles unique | Articles weighted | Authorship weight (mean) | Keywords per article (mean) | Keywords unique | Keywords weighted | Centrality | Density |
|---|---|---|---|---|---|---|---|---|---|
| 110 | | 1 | 0.5 | 47.4% | 13.0 | 0 | 0.0 | 0.0 | 0.0 |
| 111 | Geography | 0 | 0.0 | | | 12 | 0.4 | 74.1 | 0.0 |
| 112 | | 1 | 0.4 | 44.2% | 12.0 | 0 | 0.0 | 0.0 | 0.0 |
| 113 | Coronary Artery Disease | 0 | 0.0 | | | 18 | 0.4 | 162.5 | 0.0 |
| 114 | | 1 | 0.4 | 43.8% | 8.0 | 0 | 0.0 | 0.0 | 0.0 |
| 115 | | 2 | 0.4 | 21.8% | 7.7 | 0 | 0.0 | 0.0 | 0.0 |
| 116 | Population Surveillance | 0 | 0.0 | | | 3 | 0.4 | 19.8 | 0.0 |
| 117 | | 1 | 0.4 | 40.9% | 11.0 | 0 | 0.0 | 0.0 | 0.0 |
| 118 | | 1 | 0.4 | 40.9% | 5.0 | 0 | 0.0 | 0.0 | 0.0 |
| 119 | | 1 | 0.4 | 40.9% | 7.0 | 0 | 0.0 | 0.0 | 0.0 |
| 120 | | 1 | 0.4 | 40.9% | 4.0 | 0 | 0.0 | 0.0 | 0.0 |
| 121 | | 1 | 0.4 | 40.9% | 8.0 | 0 | 0.0 | 0.0 | 0.0 |
| 122 | | 1 | 0.4 | 40.9% | 8.0 | 0 | 0.0 | 0.0 | 0.0 |
| 123 | | 1 | 0.4 | 40.9% | 8.0 | 0 | 0.0 | 0.0 | 0.0 |
| 124 | | 1 | 0.4 | 40.9% | 22.0 | 0 | 0.0 | 0.0 | 0.0 |
| 125 | | 1 | 0.4 | 40.9% | 9.0 | 0 | 0.0 | 0.0 | 0.0 |
| 126 | | 1 | 0.4 | 40.9% | 7.0 | 0 | 0.0 | 0.0 | 0.0 |
| 127 | | 1 | 0.4 | 40.9% | 14.0 | 0 | 0.0 | 0.0 | 0.0 |
| 128 | | 1 | 0.4 | 40.9% | 5.0 | 0 | 0.0 | 0.0 | 0.0 |
| 129 | | 1 | 0.4 | 40.9% | 9.0 | 0 | 0.0 | 0.0 | 0.0 |
| 130 | | 1 | 0.4 | 40.9% | 9.0 | 0 | 0.0 | 0.0 | 0.0 |
| 131 | | 1 | 0.4 | 40.9% | 7.0 | 0 | 0.0 | 0.0 | 0.0 |

| ID | Name * | Articles | | Authorship weight (mean) | Keywords per article (mean) | Keywords | | Centrality | Density |
|---|---|---|---|---|---|---|---|---|---|
| | | unique | weighted | | | unique | weighted | | |
| 132 | | 1 | 0.4 | 40.9% | 12.0 | 0 | 0.0 | 0.0 | 0.0 |
| 133 | | 1 | 0.4 | 40.8% | 9.0 | 0 | 0.0 | 0.0 | 0.0 |
| 134 | Rotavirus Vaccines | 0 | 0.0 | | | 9 | 0.4 | 27.1 | 0.0 |
| 135 | Social Medicine | 0 | 0.0 | | | 3 | 0.4 | 10.5 | 0.0 |
| 136 | Critical Care | 0 | 0.0 | | | 8 | 0.4 | 56.5 | 0.0 |
| 137 | | 1 | 0.4 | 36.0% | 12.0 | 0 | 0.0 | 0.0 | 0.0 |
| 138 | | 1 | 0.4 | 36.0% | 16.0 | 0 | 0.0 | 0.0 | 0.0 |
| 139 | | 1 | 0.4 | 36.0% | 10.0 | 0 | 0.0 | 0.0 | 0.0 |
| 140 | | 1 | 0.4 | 36.0% | 5.0 | 0 | 0.0 | 0.0 | 0.0 |
| 141 | | 1 | 0.4 | 36.0% | 15.0 | 0 | 0.0 | 0.0 | 0.0 |
| 142 | | 1 | 0.4 | 36.0% | 11.0 | 0 | 0.0 | 0.0 | 0.0 |
| 143 | | 1 | 0.4 | 36.0% | 14.0 | 0 | 0.0 | 0.0 | 0.0 |
| 144 | | 1 | 0.4 | 36.0% | 4.0 | 0 | 0.0 | 0.0 | 0.0 |
| 145 | | 1 | 0.4 | 36.0% | 6.0 | 0 | 0.0 | 0.0 | 0.0 |
| 146 | | 1 | 0.4 | 36.0% | 12.0 | 0 | 0.0 | 0.0 | 0.0 |
| 147 | | 1 | 0.4 | 36.0% | 15.0 | 0 | 0.0 | 0.0 | 0.0 |
| 148 | | 1 | 0.4 | 36.0% | 16.0 | 0 | 0.0 | 0.0 | 0.0 |
| 149 | | 1 | 0.4 | 36.0% | 4.0 | 0 | 0.0 | 0.0 | 0.0 |
| 150 | Clinical Protocols | 0 | 0.0 | | | 9 | 0.4 | 93.9 | 0.0 |
| 151 | Bariatric Surgery | 0 | 0.0 | | | 13 | 0.4 | 139.0 | 0.0 |
| 152 | Annexins | 1 | 0.3 | 27.6% | 14.0 | 4 | 0.1 | 3.7 | 5.7 |
| 153 | Sleep | 0 | 0.0 | | | 14 | 0.4 | 99.5 | 0.0 |

| ID | Name * | Articles | | Authorship | Keywords per | Keywords | | Centrality | Density |
|---|---|---|---|---|---|---|---|---|---|
| | | unique | weighted | weight (mean) | article (mean) | unique | weighted | | |
| 154 | | 1 | 0.4 | 35.4% | 17.0 | 0 | 0.0 | 0.0 | 0.0 |
| 155 | Renal Dialysis | 0 | 0.0 | | | 17 | 0.3 | 150.3 | 0.0 |
| 156 | | 1 | 0.3 | 32.8% | 7.0 | 0 | 0.0 | 0.0 | 0.0 |
| 157 | | 1 | 0.3 | 32.8% | 14.0 | 0 | 0.0 | 0.0 | 0.0 |
| 158 | | 1 | 0.3 | 32.8% | 9.0 | 0 | 0.0 | 0.0 | 0.0 |
| 159 | | 1 | 0.3 | 32.8% | 18.0 | 0 | 0.0 | 0.0 | 0.0 |
| 160 | | 1 | 0.3 | 32.8% | 8.0 | 0 | 0.0 | 0.0 | 0.0 |
| 161 | | 1 | 0.3 | 32.8% | 10.0 | 0 | 0.0 | 0.0 | 0.0 |
| 162 | | 1 | 0.3 | 32.8% | 18.0 | 0 | 0.0 | 0.0 | 0.0 |
| 163 | | 1 | 0.3 | 32.8% | 13.0 | 0 | 0.0 | 0.0 | 0.0 |
| 164 | | 1 | 0.3 | 32.8% | 13.0 | 0 | 0.0 | 0.0 | 0.0 |
| 165 | | 1 | 0.3 | 30.9% | 16.0 | 0 | 0.0 | 0.0 | 0.0 |
| 166 | | 1 | 0.3 | 30.6% | 13.0 | 0 | 0.0 | 0.0 | 0.0 |
| 167 | Prisoners | 0 | 0.0 | | | 5 | 0.3 | 27.3 | 0.0 |
| 168 | | 1 | 0.3 | 28.9% | 10.0 | 0 | 0.0 | 0.0 | 0.0 |
| 169 | | 1 | 0.3 | 28.9% | 11.0 | 0 | 0.0 | 0.0 | 0.0 |
| 170 | | 1 | 0.3 | 28.9% | 13.0 | 0 | 0.0 | 0.0 | 0.0 |
| 171 | | 1 | 0.3 | 28.9% | 14.0 | 0 | 0.0 | 0.0 | 0.0 |
| 172 | | 1 | 0.3 | 28.9% | 13.0 | 0 | 0.0 | 0.0 | 0.0 |
| 173 | | 1 | 0.3 | 28.9% | 15.0 | 0 | 0.0 | 0.0 | 0.0 |
| 174 | | 1 | 0.3 | 28.9% | 13.0 | 0 | 0.0 | 0.0 | 0.0 |
| 175 | Air Pollutants, Occupational | 0 | 0.0 | | | 18 | 0.3 | 112.1 | 0.0 |

| ID | Name * | Articles unique | Articles weighted | Authorship weight (mean) | Keywords per article (mean) | Keywords unique | Keywords weighted | Centrality | Density |
|---|---|---|---|---|---|---|---|---|---|
| 176 | | 1 | 0.3 | 27.6% | 13.0 | 0 | 0.0 | 0.0 | 0.0 |
| 177 | | 1 | 0.3 | 26.5% | 18.0 | 0 | 0.0 | 0.0 | 0.0 |
| 178 | | 1 | 0.3 | 26.5% | 9.0 | 0 | 0.0 | 0.0 | 0.0 |
| 179 | | 1 | 0.3 | 25.6% | 11.0 | 0 | 0.0 | 0.0 | 0.0 |
| 180 | | 2 | 0.2 | 12.5% | 8.7 | 0 | 0.0 | 0.0 | 0.0 |
| 181 | | 1 | 0.2 | 24.8% | 14.0 | 0 | 0.0 | 0.0 | 0.0 |
| 182 | Meditation | 0 | 0.0 | | | 6 | 0.2 | 42.5 | 0.0 |
| 183 | | 1 | 0.2 | 24.2% | 21.0 | 0 | 0.0 | 0.0 | 0.0 |
| 184 | | 1 | 0.2 | 24.2% | 6.0 | 0 | 0.0 | 0.0 | 0.0 |
| 185 | Seroepidemiologic Studies | 0 | 0.0 | | | 11 | 0.2 | 76.2 | 0.0 |
| 186 | | 1 | 0.2 | 23.1% | 14.0 | 0 | 0.0 | 0.0 | 0.0 |
| 187 | Nitroimidazoles | 0 | 0.0 | | | 12 | 0.2 | 66.3 | 0.0 |
| 188 | Endemic Diseases | 0 | 0.0 | | | 5 | 0.2 | 27.6 | 0.0 |
| 189 | Health Human Resource Evaluation | 0 | 0.0 | | | 2 | 0.2 | 5.3 | 0.0 |
| 190 | | 1 | 0.2 | 18.2% | 12.0 | 0 | 0.0 | 0.0 | 0.0 |
| 191 | | 1 | 0.2 | 18.2% | 11.0 | 0 | 0.0 | 0.0 | 0.0 |
| 192 | | 1 | 0.2 | 18.2% | 16.0 | 0 | 0.0 | 0.0 | 0.0 |
| 193 | Herpes Zoster | 0 | 0.0 | | | 5 | 0.2 | 9.8 | 0.0 |
| 194 | Lumbosacral Region | 1 | 0.1 | 12.0% | 7.0 | 3 | 0.1 | 13.9 | 10.7 |
| 195 | | 1 | 0.2 | 16.0% | 10.0 | 0 | 0.0 | 0.0 | 0.0 |
| 196 | | 1 | 0.2 | 16.0% | 8.0 | 0 | 0.0 | 0.0 | 0.0 |
| 197 | | 1 | 0.2 | 16.0% | 11.0 | 0 | 0.0 | 0.0 | 0.0 |

| ID | Name * | Articles unique | Articles weighted | Authorship weight (mean) | Keywords per article (mean) | Keywords unique | Keywords weighted | Centrality | Density |
|---|---|---|---|---|---|---|---|---|---|
| 198 | | 1 | 0.2 | 16.0% | 9.0 | 0 | 0.0 | 0.0 | 0.0 |
| 199 | | 1 | 0.1 | 15.0% | 17.0 | 0 | 0.0 | 0.0 | 0.0 |
| 200 | | 1 | 0.1 | 14.6% | 16.0 | 0 | 0.0 | 0.0 | 0.0 |
| 201 | | 1 | 0.1 | 14.6% | 14.0 | 0 | 0.0 | 0.0 | 0.0 |
| 202 | | 1 | 0.1 | 14.6% | 8.0 | 0 | 0.0 | 0.0 | 0.0 |
| 203 | | 1 | 0.1 | 14.6% | 11.0 | 0 | 0.0 | 0.0 | 0.0 |
| 204 | | 1 | 0.1 | 12.9% | 15.0 | 0 | 0.0 | 0.0 | 0.0 |
| 205 | | 1 | 0.1 | 12.9% | 12.0 | 0 | 0.0 | 0.0 | 0.0 |
| 206 | Nursing | 0 | 0.0 | | | 3 | 0.1 | 7.3 | 0.0 |
| 207 | | 1 | 0.1 | 12.3% | 3.0 | 0 | 0.0 | 0.0 | 0.0 |
| 208 | | 1 | 0.1 | 12.3% | 17.0 | 0 | 0.0 | 0.0 | 0.0 |
| 209 | | 1 | 0.1 | 12.0% | 11.0 | 0 | 0.0 | 0.0 | 0.0 |
| 210 | | 1 | 0.1 | 12.0% | 15.0 | 0 | 0.0 | 0.0 | 0.0 |
| 211 | | 1 | 0.1 | 12.0% | 7.0 | 0 | 0.0 | 0.0 | 0.0 |
| 212 | | 1 | 0.1 | 12.0% | 9.0 | 0 | 0.0 | 0.0 | 0.0 |
| 213 | | 1 | 0.1 | 11.8% | 14.0 | 0 | 0.0 | 0.0 | 0.0 |
| 214 | Social Work | 0 | 0.0 | | | 2 | 0.1 | 4.5 | 0.0 |
| 215 | Sedentary Behavior | 0 | 0.0 | | | 2 | 0.1 | 8.9 | 0.0 |
| 216 | | 1 | 0.1 | 10.9% | 7.0 | 0 | 0.0 | 0.0 | 0.0 |
| 217 | | 1 | 0.1 | 10.9% | 11.0 | 0 | 0.0 | 0.0 | 0.0 |
| 218 | | 1 | 0.1 | 10.9% | 10.0 | 0 | 0.0 | 0.0 | 0.0 |
| 219 | | 1 | 0.1 | 10.9% | 14.0 | 0 | 0.0 | 0.0 | 0.0 |

| ID | Name * | Articles | | Authorship weight (mean) | Keywords per article (mean) | Keywords | | Centrality | Density |
|---|---|---|---|---|---|---|---|---|---|
| | | unique | weighted | | | unique | weighted | | |
| 220 | Social Participation | 0 | 0.0 | | | 2 | 0.1 | 4.8 | 0.0 |
| 221 | Environmental Pollution | 0 | 0.0 | | | 4 | 0.1 | 18.8 | 0.0 |
| 222 | Diseases Registries | 0 | 0.0 | | | 1 | 0.1 | 0.3 | 0.0 |
| 223 | | 1 | 0.1 | 10.2% | 8.0 | 0 | 0.0 | 0.0 | 0.0 |
| 224 | Esophageal Motility Disorders | 0 | 0.0 | | | 3 | 0.1 | 3.8 | 0.0 |
| 225 | | 1 | 0.1 | 9.6% | 10.0 | 0 | 0.0 | 0.0 | 0.0 |
| 226 | | 1 | 0.1 | 9.6% | 18.0 | 0 | 0.0 | 0.0 | 0.0 |
| 227 | | 1 | 0.1 | 9.6% | 9.0 | 0 | 0.0 | 0.0 | 0.0 |
| 228 | Intimate Partner Violence | 0 | 0.0 | | | 2 | 0.1 | 10.2 | 0.0 |
| 229 | | 1 | 0.1 | 9.2% | 10.0 | 0 | 0.0 | 0.0 | 0.0 |
| 230 | | 1 | 0.1 | 9.2% | 16.0 | 0 | 0.0 | 0.0 | 0.0 |
| 231 | | 1 | 0.1 | 8.8% | 13.0 | 0 | 0.0 | 0.0 | 0.0 |
| 232 | | 1 | 0.1 | 8.8% | 13.0 | 0 | 0.0 | 0.0 | 0.0 |
| 233 | | 1 | 0.1 | 8.8% | 14.0 | 0 | 0.0 | 0.0 | 0.0 |
| 234 | | 1 | 0.1 | 8.8% | 5.0 | 0 | 0.0 | 0.0 | 0.0 |
| 235 | | 1 | 0.1 | 8.8% | 14.0 | 0 | 0.0 | 0.0 | 0.0 |
| 236 | | 1 | 0.1 | 8.2% | 11.0 | 0 | 0.0 | 0.0 | 0.0 |
| 237 | | 1 | 0.1 | 8.2% | 11.0 | 0 | 0.0 | 0.0 | 0.0 |
| 238 | | 1 | 0.1 | 7.7% | 12.0 | 0 | 0.0 | 0.0 | 0.0 |
| 239 | | 1 | 0.1 | 7.7% | 11.0 | 0 | 0.0 | 0.0 | 0.0 |
| 240 | | 1 | 0.1 | 7.7% | 12.0 | 0 | 0.0 | 0.0 | 0.0 |
| 241 | | 1 | 0.1 | 7.7% | 6.0 | 0 | 0.0 | 0.0 | 0.0 |

| ID | Name * | Articles | | Authorship weight (mean) | Keywords per article (mean) | Keywords | | Centrality | Density |
|---|---|---|---|---|---|---|---|---|---|
| | | unique | weighted | | | unique | weighted | | |
| 242 | Antipsychotic Agents | 0 | 0.0 | | | 5 | 0.1 | 15.2 | 0.0 |
| 243 | | 1 | 0.1 | 7.4% | 12.0 | 0 | 0.0 | 0.0 | 0.0 |
| 244 | | 1 | 0.1 | 7.3% | 20.0 | 0 | 0.0 | 0.0 | 0.0 |
| 245 | | 1 | 0.1 | 7.3% | 7.0 | 0 | 0.0 | 0.0 | 0.0 |
| 246 | | 1 | 0.1 | 6.8% | 12.0 | 0 | 0.0 | 0.0 | 0.0 |
| 247 | | 1 | 0.1 | 6.4% | 9.0 | 0 | 0.0 | 0.0 | 0.0 |
| 248 | | 1 | 0.1 | 6.3% | 11.0 | 0 | 0.0 | 0.0 | 0.0 |
| 249 | Leptospirosis | 0 | 0.0 | | | 1 | 0.1 | 0.3 | 0.0 |
| 250 | | 1 | 0.1 | 5.9% | 18.0 | 0 | 0.0 | 0.0 | 0.0 |
| 251 | Anti-HIV Agents | 0 | 0.0 | | | 2 | 0.1 | 18.2 | 0.0 |
| 252 | | 1 | 0.1 | 5.5% | 7.0 | 0 | 0.0 | 0.0 | 0.0 |
| 253 | | 1 | 0.1 | 5.5% | 13.0 | 0 | 0.0 | 0.0 | 0.0 |
| 254 | | 1 | 0.1 | 5.5% | 9.0 | 0 | 0.0 | 0.0 | 0.0 |
| 255 | | 1 | 0.1 | 5.5% | 9.0 | 0 | 0.0 | 0.0 | 0.0 |
| 256 | Cholecystitis | 0 | 0.0 | | | 2 | 0.0 | 1.9 | 0.0 |
| 257 | | 1 | 0.0 | 4.6% | 13.0 | 0 | 0.0 | 0.0 | 0.0 |
| 258 | Botulism | 0 | 0.0 | | | 2 | 0.0 | 0.3 | 0.0 |
| 259 | Jejunoileal Bypass | 0 | 0.0 | | | 1 | 0.0 | 7.7 | 0.0 |
| 260 | | 1 | 0.0 | 3.9% | 15.0 | 0 | 0.0 | 0.0 | 0.0 |
| 261 | | 1 | 0.0 | 3.8% | 15.0 | 0 | 0.0 | 0.0 | 0.0 |
| 262 | | 1 | 0.0 | 3.7% | 14.0 | 0 | 0.0 | 0.0 | 0.0 |
| 263 | Cesium Radioisotopes | 0 | 0.0 | | | 2 | 0.0 | 1.7 | 0.0 |

| ID | Name * | Articles unique | Articles weighted | Authorship weight (mean) | Keywords per article (mean) | Keywords unique | Keywords weighted | Centrality | Density |
|---|---|---|---|---|---|---|---|---|---|
| 264 | Geographic Mapping | 0 | 0.0 | | | 1 | 0.0 | 1.5 | 0.0 |
| 265 | Anemia, Sickle Cell | 0 | 0.0 | | | 2 | 0.0 | 5.1 | 0.0 |
| 266 | Consensus Development Conference | 0 | 0.0 | | | 1 | 0.0 | 1.1 | 0.0 |
| 267 | Weight Perception | 0 | 0.0 | | | 1 | 0.0 | 3.2 | 0.0 |
| 268 | Deltaretrovirus Infections | 0 | 0.0 | | | 1 | 0.0 | 1.0 | 0.0 |
| | **Total** | 1903 | 576.5 | 30.3% | 8.9 | 3222 | 576.5 | | 37.1 |

Authorship weight is the share of family and community physicians in Brazil among authors, with harmonic authorship counting. Authorship weighing is used for the “weighted” number of articles and keywords, as well as for the mean number of keywords per article. * Research themes without a name are those with no keywords.

*Suppl. Table 2 – Postgraduate trajectories of family and community physicians leading to articles, by research theme*

| **Characteristic** | **Humans** | | **Primary Health Care** | | **Animals** | | **Total** | |
|---|---|---|---|---|---|---|---|---|
| Gender * | | | | | | | | |
| Female | 95.2 | 35.3% | 59.7 | 31.1% | 16.5 | 60.7% | 205.3 | 35.8% |
| Male | 174.7 | 64.7% | 132.2 | 68.9% | 10.7 | 39.3% | 367.6 | 64.2% |
| Mode of specialization | | | | | | | | |
| Certification | 70.5 | 26.0% | 61.7 | 31.7% | 11.7 | 42.9% | 169.8 | 29.4% |
| Medical residency | 200.6 | 74.0% | 132.9 | 68.3% | 15.5 | 57.1% | 407.7 | 70.6% |
| Knowledge area | | | | | | | | |
| None | 39.9 | 14.7% | 37.9 | 19.5% | 2.6 | 9.6% | 97.7 | 16.9% |
| Medicine | 110.4 | 40.7% | 26.6 | 13.7% | 10.0 | 36.7% | 179.2 | 31.0% |
| Collective health | 81.7 | 30.1% | 80.2 | 41.2% | 2.4 | 8.7% | 191.3 | 33.1% |
| Other | 39.1 | 14.4% | 49.9 | 25.6% | 12.2 | 45.0% | 109.3 | 18.9% |
| Postgraduate degree | | | | | | | | |
| None | 39.9 | 14.7% | 37.9 | 19.5% | 2.6 | 9.6% | 97.7 | 16.9% |
| Master's | 75.6 | 27.9% | 80.6 | 41.4% | 3.3 | 12.0% | 183.9 | 31.8% |
| PhD | 155.6 | 57.4% | 76.1 | 39.1% | 21.3 | 78.4% | 296.0 | 51.3% |
| Year of publication | | | | | | | | |
| 1985–1998 | 16.4 | 6.1% | 6.2 | 3.2% | 0.8 | 2.8% | 28.8 | 5.0% |
| 1999–2003 | 17.1 | 6.3% | 1.9 | 1.0% | 1.3 | 4.8% | 23.8 | 4.1% |
| 2004–2008 | 50.3 | 18.6% | 25.9 | 13.3% | 6.2 | 22.9% | 94.8 | 16.4% |
| 2009–2013 | 80.8 | 29.8% | 77.8 | 40.0% | 5.6 | 20.5% | 191.2 | 33.1% |
| 2014–2018 | 106.4 | 39.3% | 82.8 | 42.6% | 13.3 | 49.1% | 239.0 | 41.4% |

Frequencies are weighted by harmonic authorship counting. * It was not possible to infer the gender of 16 family and community physicians, who authored 4.5 (0.8%) articles with weighing.
